# Common Electrophysiology Biomarkers Collected at Home Robustly Track Depression Recovery With Deep Brain Stimulation

**DOI:** 10.64898/2026.04.13.26350107

**Authors:** Elif Ceren Fitoz, Sankaraleengam Alagapan, Jungho Cha, Ki Sueng Choi, Martijn Figee, Brian H. Kopell, Mosadoluwa Obatusin, Stephen Heisig, Tanya Nauvel, Aida Razavilar, Parisa Sarikhani, Isha Trivedi, Jaimie Gowatsky, Jessa Alexander, Romain Guignon, Maryam Khalid, Gutemberg Bobby Forestal, Ha Neul Song, Tim Denison, Shannon O’Neill, Shreesh Karjagi, Allison C. Waters, Patricio Riva-Posse, Helen S. Mayberg, Christopher J. Rozell

## Abstract

Subcallosal cingulate cortex (SCC) deep brain stimulation (DBS) can provide relief for individuals with Treatment Resistant Depression (TRD), but ongoing clinical management remains challenging due to nonspecific symptom fluctuations that can obscure core depression recovery on standard rating scales. Objective, stable biomarkers that selectively track the therapeutic effects of SCC DBS are therefore essential for developing principled decision support systems to guide stimulation adjustments. Recent bidirectional DBS systems enable chronic recording of local field potentials (LFPs) and prior work using the Activa PC+S device identified an electrophysiological signature of stable clinical recovery. However, translation to practical clinical deployment requires demonstrating that this biomarker is robustly generalizable, specific to the impact of the DBS therapy, and deployable in real-world recording contexts. To address this need, we developed an at-home SCC LFP data collection platform (built on the Medtronic Summit RC+S system) enabling at home data collection for a new cohort of ten SCC DBS participants with TRD (ClinicalTrials.gov identifier NCT04106466). Using longitudinal LFP recordings collected from this system, we report findings demonstrating that the previously reported biomarker of stable recovery generalizes across subject cohorts and devices without any model retraining, is robust to common potential confounds (including time of day and stimulation status), and shows symptom specificity, sensitivity and stability necessary to support clinical decision making. Across both cohorts, biomarker changes show relationships to pre-DBS white matter structure and network function measured using diffusion MRI and resting-state functional MRI (rsfMRI). These findings replicating and extending previous results support the biomarker’s utility as a foundation for scalable, electrophysiology-informed decision support in SCC DBS.

## INTRODUCTION

Deep brain stimulation (DBS) of the subcallosal cingulate cortex (SCC) can provide relief for individuals with Treatment Resistant Depression (TRD), with response rates as high as 90% in open-label studies (Mayberg et al. 2005; Holtzheimer et al. 2012; Puigdemont et al. 2012; Lozano et al. 2008; Ramasubbu et al. 2018; Riva-Posse et al. 2018; Alagapan et al. 2023) and longitudinal evidence demonstrating long-term therapeutic effectiveness (Crowell et al. 2019; Holtzheimer et al. 2017; Kennedy et al. 2011; Puigdemont et al. 2012; Ramasubbu et al. 2020; Merkl et al. 2018; Himes et al. 2025).

Precision connectomics-based targeting strategies have recently been shown to improve treatment response (Riva-Posse et al. 2014, 2018; Ramasubbu et al. 2020; Alagapan et al. 2023; Himes et al. 2025), with large pooled datasets linking therapeutic nonresponse to insufficient engagement of a common treatment network (Riva-Posse et al. 2018). While optimized targeting has improved response rates overall, ongoing clinical management decisions of individual SCC DBS patients remain complicated by persistent nonspecific symptoms often distinct from the presenting TRD syndrome that responds to DBS, including sleep disturbances, anxiety, and individualized reactions to everyday stressors (Crowell et al. 2015). Critically, these symptoms can confound gold-standard clinical rating scales such as the Hamilton Depression Rating Scale (HDRS; (Hamilton 1960)), leaving psychiatrists without objective measures of DBS-responsive core symptom recovery state to inform stimulation adjustments. Effective decision support systems for SCC DBS would establish generalized treatment principles to reduce idiosyncratic decision making and lower the activation barriers for clinical teams to increase access (Forum on Neuroscience and Nervous System Disorders et al. 2024).

Developing principled decision support systems requires reliable measures that are stable and selective for the primary therapeutic effects of SCC DBS. Recent neurotechnology advances have enabled bidirectional DBS devices that can deliver electrical stimulation as well as capture electrophysiological activity from the targeted brain region (Stanslaski et al. 2018, 2012; Zamora et al. 2022; Goyal et al. 2021) that have been deployed in a variety of disorders (Gilron et al. 2021; Schmidt et al. 2024; Opri et al. 2020; Provenza et al. 2021, 2024; Sladky et al. 2021; Scangos et al. 2021). Our previous work used the first generation research prototype Activa PC+S (Stanslaski et al. 2012) in a cohort (N=5) of subjects to identify a common electrophysiological biomarker of stable clinical recovery (Alagapan et al. 2023) using longitudinally recorded SCC local field potentials (LFPs) downloaded during in-clinic weekly visits. This biomarker accurately tracked individual recovery trajectories, showed a predictable response to clinician initiated stimulation adjustments, and demonstrated a forewarning of impending relapse in a patient over four weeks prior to awareness of clinical symptoms. In contrast to adaptive DBS approaches that are now approved to make real-time stimulation adjustments based on biomarkers of the fast symptom fluctuations in Parkinson’s Disease (Bronte-Stewart et al. 2025), this SCC LFP biomarker demonstrates an objective brain signal that can inform clinical decision making on the time scales of recovery from chronic depression and infrequent but necessary adjustments that have been successful with continuous SCC DBS therapy.

To support the goal of developing a practical and scalable decision support system for SCC DBS that leverages an electrophysiology biomarker, we require any proposed biomarker to be generalizable across subjects, specific and sensitive to the core depression syndrome treated by SCC DBS, stable during unrelated behavioral fluctuations that can confound depression symptom rating scales, robust to recording conditions, and able to be collected with high-compliance on commercially available devices. Here we report our development of an at-home data collection system and use of that data to generalize previously reported SCC LFP biomarkers toward practical utility for decision support during ongoing DBS treatment. The data collection system is built on top of the Medtronic Summit RC+S research prototype DBS device (Stanslaski et al. 2018) to capture SCC LFPs, mood ratings, and video diaries using a mobile device (Microsoft Surface Pro). This system facilitated twice-a-day data recording in participants’ home environments with high compliance, significantly increasing the richness of collected data while using immediate downloads to a HIPAA-compliant research server to eliminate the need for frequent in-clinic visits. Using longitudinal LFP data collected in a new cohort (N=10) of individuals with TRD undergoing SCC DBS with this platform, we report findings demonstrating that the previously reported biomarker of stable recovery generalizes across subject cohorts and distinct devices without model retraining, is robust to common potential confounds (including time of day and stimulation status), and shows symptom specificity, sensitivity and stability necessary to support clinical decision making. Using this expanded population of SCC DBS patients across two cohorts with longitudinal LFP recordings, we also show consistent relationships between changes in the electrophysiology and pre-DBS imaging of white matter structure and network function using diffusion MRI and resting-state functional MRI (rsfMRI), respectively.

## RESULTS

### Platform for chronic monitoring in SCC DBS

The Summit RC+S system (Medtronic Inc, Minneapolis, USA) is an investigational, rechargeable DBS platform that is the successor to the previous generation Activa PC+S investigational device. The platform offers collection of intracranial LFP and modification of stimulation parameters through the device’s application programming interface (API). We leveraged the API to develop a custom application, the Mount Sinai Summit Home Application (MSSHA), to collect LFP, video diaries and visual analogue assessment scale (VAS) of mood, motivation, energy and alertness, and daily self-reported Computerized Adaptive Testing for Mental Health (CAT-MH; (Gibbons et al. 2012) via an external tablet (Microsoft Surface Pro, Microsoft Inc, Redmond, USA). Each session included symptom characterization (including a customized Ecological Momentary Assessment and daily online severity metrics), an 8-minute LFP recording (with stimulation ON and stimulation OFF) streamed via MICS-Telemetry (Stanslaski et al. 2018), and an open-ended video diary (Fig. 1.c.). This workflow enabled twice daily data collection (approximately 15 minutes each) in addition to a weekly examiner-rated 17-item HDRS (HDRS-17), which was a substantial improvement over data collection with the Activa PC+S requiring weekly in-clinic visits to record stimulation-off data or download stimulation-on recording snapshots collected during the week (Fig. 1.b.). While the Summit RC+S system specifics differ from the current commercially available Percept™ system, this investigational setup demonstrated that it is possible to collect high quality data with high compliance from TRD patients with at-home data collection. The present study focuses on the generalizability of the prior LFP-derived biomarker across patients and devices, while future work will examine participant-specific characterization using other data modalities (e.g., video diaries, etc.).

**Figure 1:**
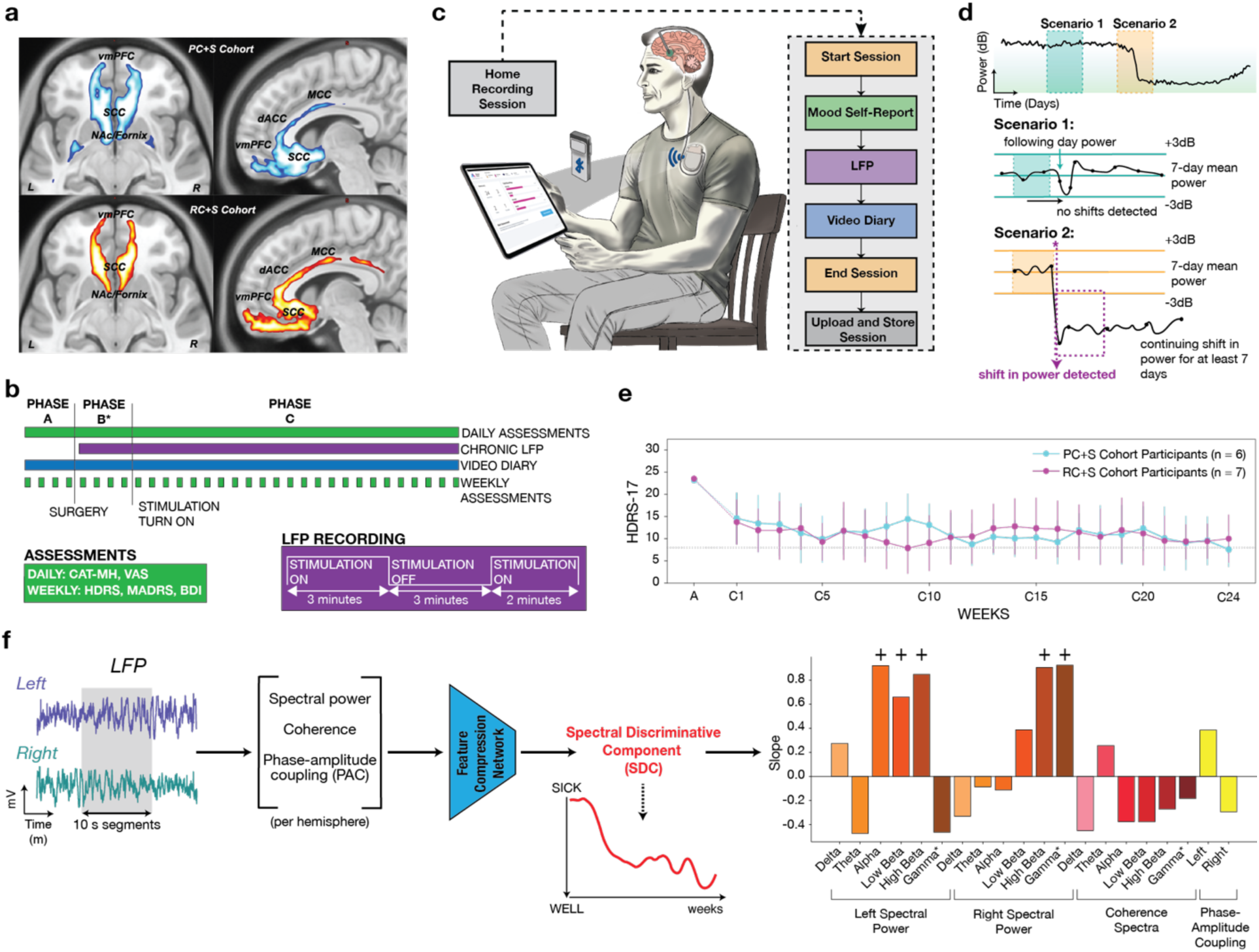
a) Coronal and sagittal sections showing consistency in the common activation maps for the prior PC+S (blue, upper) and current RC+S (orange, lower) cohorts. b) Data collection schedule for the current RC+S cohort. LFP recordings, CAT-MH and visual analog scales (VAS) collected at home daily from participants available for analysis. HDRS-17, MADRS and BDI assessed weekly during visits. c) Illustration of data collection at home using the Mt. Sinai Depression DBS Home Application d) Illustration of criteria used for the exclusion of participants whose LFP recordings exhibited unexplained changes in signal quality. A custom moving window–based shift-detection algorithm was used to identify changes in power (dB) that persisted for at least 7 days. A shift was defined as a ≥3 dB increase or decrease in the current day’s power relative to the mean power of the preceding 7-day window. e) Trajectory of HDRS-17 scores over 24 weeks for participants in both cohorts (PC+S, RC+S) whose LFP recordings were used in the study. f) Derivation of the Spectral Discriminative Component (SDC) from bilateral LFP recordings. Left- (purple) and right- (teal) hemisphere LFPs were split into 10-s segments, from which features of spectral power, inter-hemispheric coherence, and phase-amplitude coupling (PAC) were extracted and were then compressed by a feature compression network into a single scalar value called the spectral discriminative component (SDC). The SDC was tracked over weeks of recording, with higher values corresponding to the symptomatic (sick) state and lower values to the recovered (well) state. The bar plot shows the slope of each input feature along the SDC recovery axis. Positive and negative slopes indicate features that increased or decreased (respectively) as the SDC moved from the sick to the well state. The bar plot was reproduced from (Alagapan et al. 2023).

To evaluate adherence and data quality, participant compliance was quantified based on the completion of the prescribed twice-daily recordings (morning and evening) over the predefined 24-week study period. The average compliance rate across participants was 93.95% (s.d. 7.01), with subjects responding clinically to DBS having a rate of 97.28% (s.d. 2.31) and non-responding subjects having a rate of 80.65% (s.d. 1.79). While response rates are high overall, the lower response rates for non-responding subjects further indicates the value of objective biomarkers that do not rely on self-reports. Weekly compliance rates for individual participants with the cohort-level summary are presented in Supp. Fig. 3..

### Clinical outcomes of new RC+S cohort

Ten participants meeting the study eligibility criteria were implanted with the Summit RC+S DBS system. The DBS targets were determined from diffusion MRI to be at the intersection of white matter pathways for SCC DBS as described in previous studies (Riva-Posse et al. 2018; Alagapan et al. 2023) (Fig. 1.a., Supp. Fig. 1.). At the cohort level, the presurgical average HDRS-17 score was 23.93 (s.d. 2.33), which significantly decreased to 9.6 (s.d. 4.67) by the end of the 6 month observation period. Seven out of ten participants responded to the therapy (achieving a 50% reduction from the presurgical baseline at the predefined 24-week point) and four participants showed remission (HDRS-17 score less than 8). One additional participant (R002) met the response criterion at week 25 (one week beyond the predefined endpoint but prior to the discontinuation phase) and is included in the responder group in all data analyses. The first five participants in this cohort (RC+S-1) received chronic stimulation after a 4-week postsurgery recovery phase during which stimulation remained off. The subsequent five participants (RC+S-2) began chronic stimulation one day postsurgery and prior to discharge from the hospital. The two sub-cohorts were combined for analysis, as there were no differences in response rates (4 responders per group).

Chronic electrophysiological data was available for analysis for 7 participants after excluding 3 participants from the analysis due to sudden unexplained shifts in recording properties from the investigational devices (Fig. 1.d., Supp. Fig. 4.). Of the 7 participants with available electrophysiology data for analysis, there was a diversity of response characteristics: 4 responded to the treatment at the predefined 24-week, 1 responded at week 25, and 2 were deemed non-responders. The responders began with a mean HDRS-17 score of 22.65 (s.d. 1.24) and concluded the chronic stimulation period with a mean score of 7.0 (s.d. 3.29). The clinical trajectories of participants used in the study are similar to the previous PC+S cohort (Alagapan et al. 2023), shown in Fig. 1.e. and Supp. Fig. 2. Week by week comparisons of HDRS-17 scores between cohorts using a variety of metrics (including the baseline presurgery HDRS-17, the endpoint HDRS-17 at week 24, the time to stable response, and the Illness Density Index (Kelley et al. 2012) measuring aggregated recovery trajectories over time) revealed no significant difference at any week except a single isolated week (week 9; two sided Mann Whitney U test). A linear mixed effects model (HDRS-17 ∼ week × group, random intercept per subject) further showed no difference between cohorts in overall HDRS-17 level (group effect, *p* = 0.825) or in rate of recovery (group × week interaction, *p* = 0.310), while both cohorts improved significantly over time (week effect, *p* = 0.0011) (Supp. Fig. 2.).

### SDC biomarker generalizes to RC+S cohort

A central test of the clinical utility of the biomarker is whether it generalizes to a new patient population and a different recording device without any model retraining or recalibration. We therefore evaluated the model previously trained on the PC+S cohort (Alagapan et al. 2023) directly to the RC+S cohort, without retraining or recalibrating on any data from this new cohort. Briefly reviewing this previous model, LFP spectral features were used to train a neural network classifier to distinguish the ’sick’ and ’stable response’ states. We then trained a generative model to generate synthetic LFP features that were fed into the fixed classifier model. In this generative causal explainer (GCE) producing synthetic LFP features, the generator training rule encourages both the typical replication of the original data statistics as well as encouraging a dimension within the latent space to capture the composite data characteristics used by the classifier to distinguish the classes (O’Shaughnessy et al. 2020). This latent dimension is the electrophysiological biomarker for identifying stable recovery derived from the GCE model, denoted the spectral discriminative component (SDC) (Alagapan et al. 2023). The SDC therefore serves as an LFP marker of sick vs. stable recovery state, with higher values indicating the ’sick’ state and lower values indicating the ’stable response’ state (Fig. 1.f.).

Specifically, in this study we extracted spectral features from the RC+S LFP recordings collected during morning sessions with stimulation turned off. Among the seven participants observed until the 1 week discontinuation of chronic stimulation, stable response transitions were identified using the SDC (threshold of 0.5 identified from the ROC curve in PC+S cohort) and clinical measures as previously described (Alagapan et al. 2023) (i.e., crossing response threshold and never returning above threshold for more than one week). Overall, the weekly SDC biomarker-derived depression vs stable recovery states predicted the HDRS-17-derived depression vs stable recovery states in the RC+S cohort with an accuracy of 0.72 ± 0.16, which is comparable to the accuracy of prediction in the PC+S cohort (ACC: 0.86 ± 0.11; two-sided Mann–Whitney U test, *U =* 25.50, *p* = 0.22, r = −0.457, 95% CI = [−1.00, 0.23]) (Fig. 2.a,b., Supp. Fig. 6.). This generalization of the biomarker to a new cohort is a notable demonstration of robustness for several reasons: the original training set was a small sample that may have suffered from contributed to statistical overfitting, the two distinct DBS platforms used to collect data have different electronics that could have introduced a distribution shift that negatively affected the biomarker, and this cohort was implanted and treated by a different clinical team.

**Figure 2:**
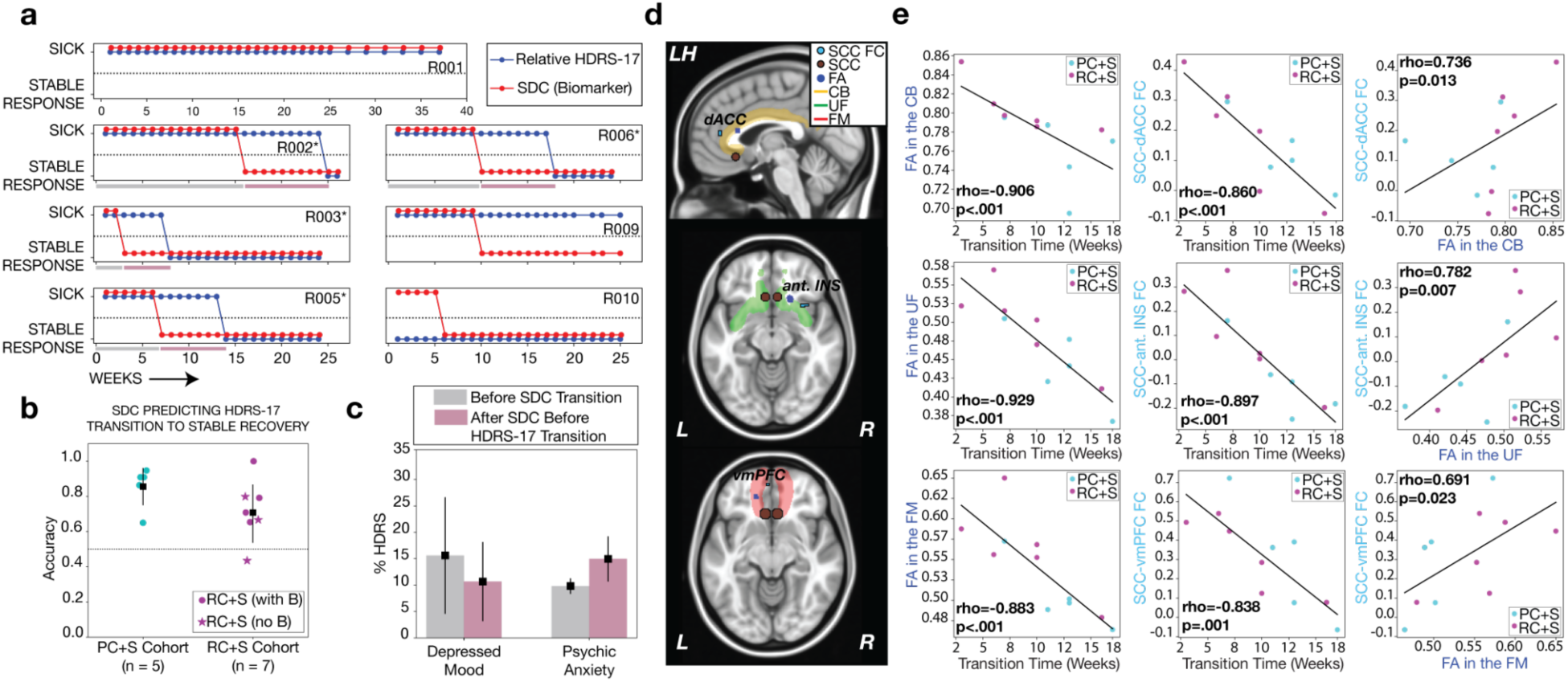
a) Trajectories of depression state inferred from HDRS-17 and SDC in the RC+S cohort. Blue lines denote the HDRS-17-defined sick/stable response states while red lines denote the SDC identified response states using the threshold of 0.5. b) Accuracy of SDC-defined states predicting HDRS-17-defined states. Markers indicate individual participants. Magenta circles indicate participants who went through a B phase (delay of chronic stimulation onset for one month postsurgery) and stars indicate participants who did not. Black squares indicate mean and error bars indicate standard deviation. c) Comparison of depressed mood and psychic anxiety sub-items contribution to total HDRS-17 in the period before SDC transition (Gray bar) and the period between SDC-defined transition and HDRS-17-defined transition (Pink bar). Black dots indicate mean across 4 participants and error bars indicate standard deviation. See Supp. Fig. 7. for individual participant plots. d) Localization of regions within the targeted network that exhibited significant correlations with SDC-defined transition weeks. Red circle indicates the SCC DBS target. Dark blue circle indicates regions where FA variations are significantly correlated with transition weeks in the target network. Light blue indicates regions where functional connectivity between SCC and target network region exhibited significant correlation with SDC-defined transition weeks. e) Correlations between SDC-defined transition weeks and FA in target network regions (first column), and functional connectivity between SCC and target network regions (second column) and correlations between FA and functional connectivity between SCC and target network regions (third column) for areas shown in 2.d.

Building on prior evidence linking white matter integrity to therapeutic responsiveness (Alagapan et al. 2023), we also find a generalization to this new cohort of robust observations of negative correlations between the time to stable response (defined by the SDC) and fractional anisotropy (FA) within white matter tracts implicated in SCC-DBS efficacy. Specifically, lower FA in the left cingulum bundle (CB; ρ = –0.91, *p* < 0.001), right uncinate fasciculus/ventral amygdalofugal pathway (UF/vAFP; ρ = –0.93, *p* < 0.001), and left forceps minor (FM; ρ = –0.88, *p* < 0.001) was associated with longer time to response (Fig. 2.d,e.). Similarly, decreased functional connectivity between the SCC and discrete cortical regions, determined via whole brain analyses independent of the WM findings—including the left dorsal anterior cingulate cortex (dACC; ρ = –0.86, *p* < 0.001), right anterior insula (ρ = –0.90, *p* < 0.001), and left ventromedial prefrontal cortex (vmPFC; ρ = –0.84, *p* = 0.001)—was also associated with delayed response. Notably, FA within these tracts was positively correlated with adjacent SCC–cortical functional connectivity (CB–SCC–dACC: ρ = 0.74, *p* = 0.013; UF/vAFP–SCC–anterior insula: ρ = 0.78, *p* = 0.007; FM–SCC–vmpf (frontal pole): ρ = 0.69, *p* = 0.023), suggesting a structure–function coupling wherein focal microstructural disruption may impede the functional integration of specific limbic network connections critical for therapeutic response (Fig. 2.e.).

### Biomarker demonstrates specificity to guide DBS adjustments

The previous sections have demonstrated that stability of the SDC accurately predicts stability of HDRS-17-defined recovery state. This concordance of biomarker and clinical endpoint measures is particularly useful for decision support, as it is well known that the HDRS-17 is unreliable as a general measure of symptom fluctuations and not specific to the type of intervention (if any) required at any given time. For example, the HDRS-17 is not always reliable as an ongoing metric of symptoms in patients who have recovered due to the fact that transient stressors may affect the HDRS-17 (interest, appetite, sleep, anxiety) even in the absence of a relapse or recurrence of a depressive episode (Bagby et al. 2004). As such, the SDC will be most valuable if it is specific to those aspects of depression that are treated by SCC DBS (potentially requiring stimulation adjustment) and invariant to transient symptom reports that may impact weekly HDRS-17 scores but may require alternative interventions (psychotherapy, medications).

To gain insight into the specificity of the SDC in real world scenarios, we examined four responding participants (R002, R003, R005, R006) where the SDC biomarker indicated a stable response distinctly before the HDRS-17 reached a stable response. In this group of participants, the period when the SDC and HDRS-17 were incongruent (i.e., the SDC indicating stable response while HDRS-17 indicated sick state) relative to the pre-response period (i.e., before the SDC indicated stable response) was marked by a relatively higher proportion of the ‘Psychic Anxiety’ subitem to the total HDRS-17 compared to the proportion of the ‘Depressed Mood’ subitem (Fig. 2.c. and Supp. Fig. 7.). We interpret this shift in sub-item composition not as a measure of anxiety as a construct, but as an indication that the residual elevation in the total HDRS-17 during this window is driven by symptoms that SCC DBS does not treat (i.e., persistent rumination and worry) rather than by the core depressed mood that does respond to stimulation. As this study is a replication of the primary LFP biomarker’s performance to capture the categorical HDRS-17 criteria for stable response, we use the psychic anxiety subitem as the available readout of these residual symptoms, rather than introducing a separate anxiety scale. This demonstration that the SDC tracks with the core depression symptoms responsive to SCC DBS extends our previous case study (Alagapan et al. 2023) and further confirms our previous clinical observations (Crowell et al. 2015) that anxiety symptoms can persist despite depression recovery, and in the absence of comorbid generalized anxiety disorder on the screening diagnostic interview. While the SDC-HDRS-17 incongruence of this subgroup decreases the overall accuracy reported in Fig. 2.b., the selectivity of the SDC for DBS-sensitive symptom fluctuations increases its additional utility to direct treatment decisions to alternatives other than DBS. As DBS was ineffective in attenuating anxiety symptoms in all cases, time courses for participants R002, R005 and R006 are detailed below.

### Biomarker demonstrates sensitivity and stability to guide timing of interventions

In addition to specificity, the biomarker will be most useful for decision support if it demonstrates both sensitivity to deteriorating DBS-responsive clinical state and stability during behavioral fluctuations that do not require DBS stimulation changes. A biomarker sensitive enough to indicate insufficient stimulation leading to impending relapse can potentially serve as an alarm indicating the need for urgent DBS adjustments before relapse is clinically apparent (as has been shown previously; (Alagapan et al. 2023). Importantly, to limit false alarms, this biomarker should also be stable in situations of transient distress or emergence of DBS-insensitive symptoms such as anxiety, sleep changes or medication side effects.

As one method for characterizing this combination of sensitivity and stability, we investigate the SDC responses to stimulation discontinuation at various stages of treatment and recovery stability. In particular, we examine the SDC time course of participant R002 when they experienced an unplanned double-blind discontinuation of stimulation at week 9. This discontinuation happened in the early phase of treatment when the biomarker was on a trajectory toward recovery but still changing significantly each week. One week after discontinuation, the SDC worsened (Fig. 3.a.) to the point of indicating relapse by week 12. Stimulation was coincidentally resumed at week 12, followed by the SDC improving one week later to the point of indicating stable recovery from week 16 onward.

**Figure 3:**
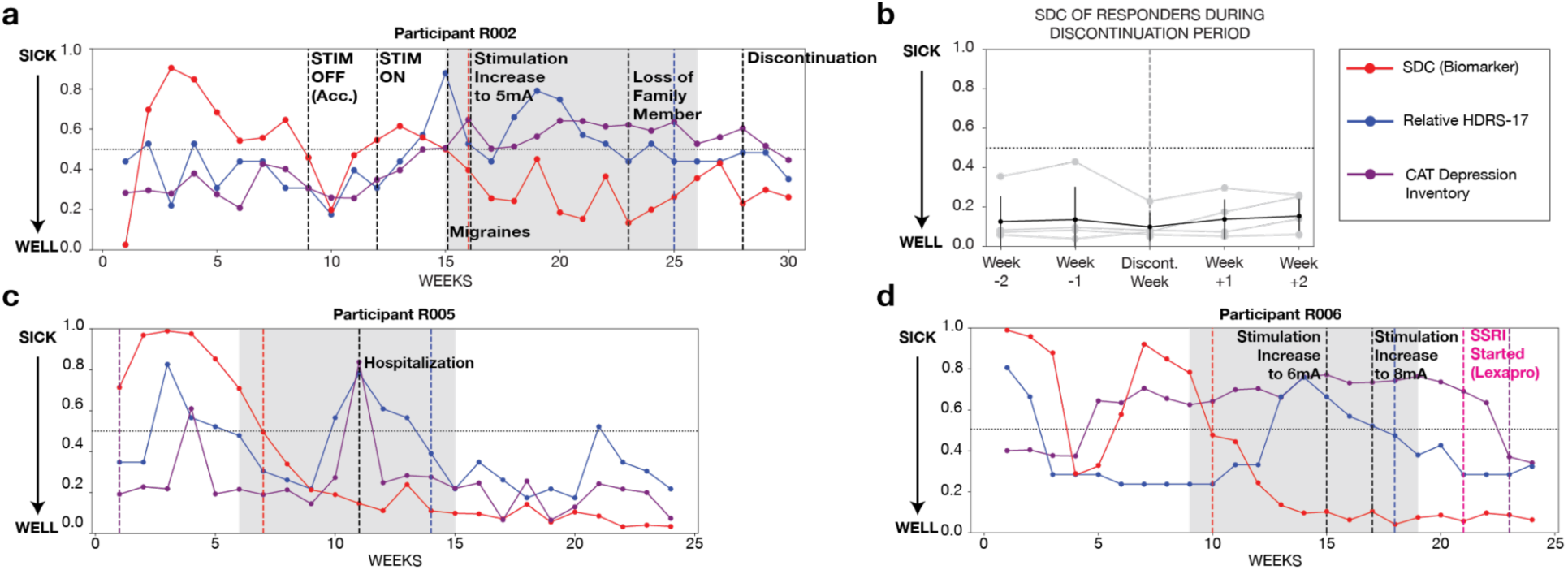
a) Trajectory of SDC (red), relative HDRS-17 (blue), and CAT-DI (purple) scores in patient R002. Black vertical dashed lines mark stimulation changes and major clinical events. An accidental double blind stimulation discontinuation at week 9 was followed by a progressive SDC worsening over the subsequent 2 weeks without stimulation, crossing the 0.5 threshold by week 12 indicating a sick state requiring intervention. After stimulation was resumed at week 12; SDC improved and remained below 0.5 from week 16 onward, showing a consistent stable response despite relative HDRS-17 remaining elevated due to symptoms unrelated to core baseline depression (migraine headaches and bereavement). A later planned 1-week discontinuation (week 28) during this established response produced no change in SDC and relative HDRS-17, indicating that the impact of stimulation withdrawal on SDC may depend on treatment stage and duration. b) SDC biomarker dynamics in responders during the stimulation discontinuation period (±2 weeks). The horizontal dotted line at 0.5 marks the SDC threshold for well (below) and sick (above) states, and the vertical dashed line indicates the week of stimulation discontinuation. In RC+S responders (n = 5), SDC remains below threshold and unchanged across the 1-week stimulation-off period, indicating a stable well-state that is not perturbed by brief stimulation withdrawal. c) Trajectory of SDC (red line), relative HDRS-17 (Blue line), and CAT Depression Inventory (CAT-DI) observed in participant R005. The period between when the SDC indicated stable response (red vertical dashed line) and HDRS-17 indicated stable response (blue vertical dashed line) included a hospitalization (black vertical dashed line) which was concurrent with an increase in relative HDRS-17 and CAT-DI. SDC predicted stable recovery of core depression, which is consistent with a return to therapeutic response without stimulation change. d) Trajectory of SDC (red line), relative HDRS-17 (Blue line), and CAT Depression Inventory (CAT-DI) observed in participant R006. The period between when the SDC indicated stable response (red vertical dashed line) and HDRS-17 indicated stable response (blue vertical dashed line) included an increase in relative HDRS-17 and adjustments in stimulation (black vertical dashed lines). While stimulation amplitude changes were followed by decrease in relative HDRS-17, SDC predicted stable recovery and CAT-DI did not exhibit any changes until a pharmacological intervention (SSRI) was introduced (pink vertical dashed line).

In contrast, when the same participant underwent a planned 1-week discontinuation after 6 months of sustained treatment (week 28), SDC and HDRS-17 remained stable (Fig. 3.a.). This pattern was consistent across other responders, who each underwent a planned 1-week blinded discontinuation after approximately 6 months of chronic stimulation. The SDC remained stable in all cases (Fig. 3.b.), although HDRS-17 changes were heterogenous across individuals. While most participants were clinically stable (similar to the previous PC+S cohort undergoing a 1-week discontinuation; unpublished data), two subjects exhibited brief symptom recurrence during the discontinuation week before returning to baseline within the observation window (Supp. Fig. 11.). This stage-dependent behavior suggests SDC could serve different purposes in the early and later stages of ongoing DBS treatment. As another method for characterizing this combination of sensitivity and stability, we highlight two participants from the RC+S cohort where clinical instabilities occurred during ongoing stimulation that were (in hindsight) unrelated to depression relapse and did not require stimulation adjustment. Participant R005 began as a typical responder to SCC DBS but exhibited an episode of rapid HDRS-17 worsening during a self-reported life stressor (blue line in Fig. 3.c.) leading to a self-admitted hospitalization (Week 11, solid vertical line in Fig. 3.c.). While the HDRS-17 change during this period suggested a relapse in depression, SDC indicated that the participant was in a stable response state, which in turn suggested that changes in DBS may not be effective in reducing the new symptoms. This conclusion was validated when the clinical decision was made to address sources of distress adjunctively without an increase in DBS stimulation dose and the symptom severity decreased the following weeks with supportive psychotherapy without any change in stimulation or adjunct medication.

Participant R006 was on no medication at the time of implant and exhibited a rapid response in HDRS-17 only two weeks after commencing SCC DBS stimulation (Fig. 3.d.). This participant then had emergent ruminative symptoms that resulted in a gradual increased HDRS-17 beginning around week 11. In contrast, the SDC indicated a transition to stable response at Week 10 with little fluctuation associated with the behavioral instability. While the SDC indicating stable response implied stimulation change would be ineffective, the clinical decision was made (without knowledge of the SDC status) to increase stimulation amplitude in an attempt to resolve the reported distressing rumination. Two stimulation amplitude increases (to 6 mA at Week 15 and 8 mA at Week 17) modestly lowered the HDRS-17 (due to relatively minor psychomotor improvements) but did not change the participant’s chief symptoms, corroborated by the CAT-Depression Inventory (CAT-DI), the depression severity scale within the CAT-MH, remaining high. The symptoms were finally addressed by the reintroduction of a selective serotonin reuptake inhibitor (SSRI) at week 21, which resulted in a relatively rapid improvement in subjectively-reported symptoms and CAT-DI scores. Notably, the patient had discontinued this class of medication due to loss of effectiveness and side effects long before DBS implantation. The ineffective stimulation adjustment in this case (followed by response to an adjunctive therapy) supports the notion that the SDC can be stable despite concurrent clinical fluctuations that require alternative treatments.

### Biomarker demonstrates recording robustness to enable reliable data collection

Another important consideration for biomarkers used in decision support systems is their robustness to common confounding variables that could affect recording, including stimulation status and time of day. The SDC was developed using data collected with stimulation-off data due to stimulation-induced recording artifacts at the sensing electrodes in Activa PC+S’s LFP measurement (Tiruvadi et al. 2022) that corrupted the accurate computation of spectral features like coherence. The Summit RC+S system improves robustness to stimulation artifacts (Stanslaski et al. 2018) thereby enabling the computation of SDC during ongoing stimulation. However, it is unknown if the electrophysiology dynamics underlying the SDC substantially change during stimulation, potentially rendering the SDC unreliable using stimulation-on data. To verify if SDC computed with stimulation-on data was effective as a biomarker, we compared it against SDC computed with stimulation-off data both collected within each recording session and compared SDC-derived recovery states with HDRS-17-derived states as before.

Comparisons showed that SDC-ON predictions of stable response were highly correlated with the default SDC in participants (Fig. 4.a., Supp. Fig. 8.) and similarly demonstrated high accuracy in predicting HDRS-17 states (ACC: 0.83 ± 0.09, n = 6 participants; two-sided Wilcoxon signed-rank test, W = 1.0, *p* = 0.125, r = 0.63, 95% CI = [−0.369, 0.020]) (Fig. 4.b.). This allows transition to future acquisitions without requiring discontinuation of treatment, potentially reducing the need for specialized hardware capabilities, shortening collection times, and increasing compliance.

**Figure 4:**
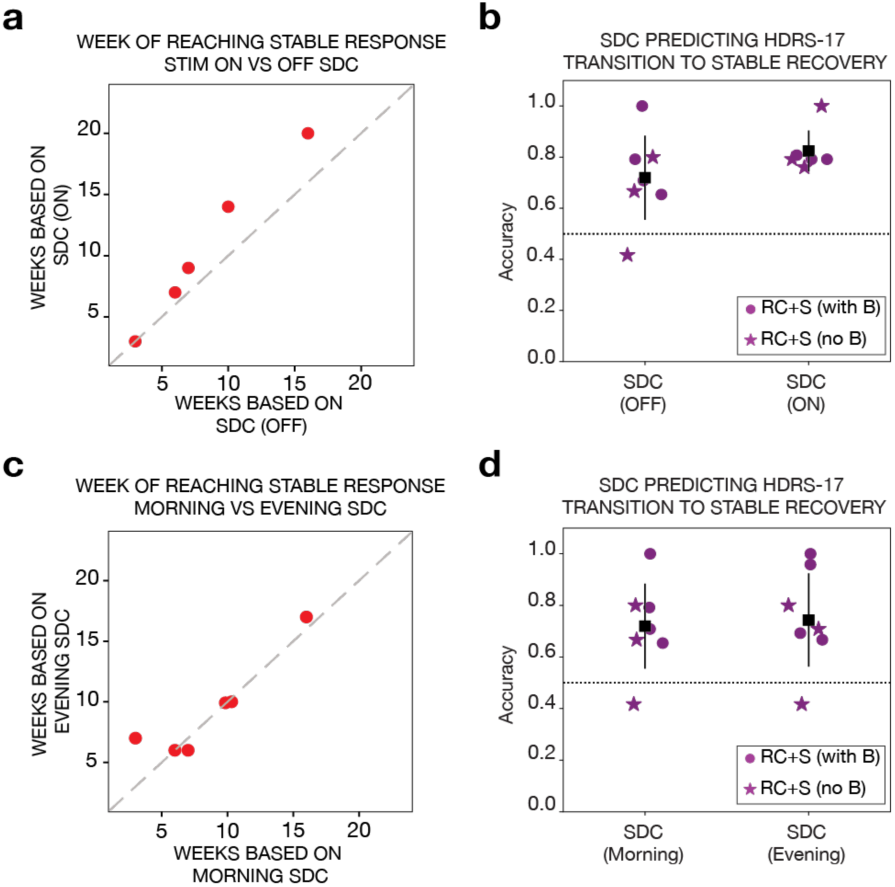
a) Correlation of stable response weeks inferred with using the standard SDC and SDC obtained using the recordings during stimulation. b) Accuracy summary for stimulation off and on biomarkers in predicting HDRS-17 transition to stable response. c) Correlation of stable response weeks inferred from SDC using the morning and SDC using the evening recordings. d) Accuracy summary for morning and evening biomarkers in predicting HDRS-17 transition to stable response.

Furthermore, given recent observations on circadian rhythm related changes in LFP (van Rheede et al. 2022, 2024; Provenza et al. 2024), we also tested if the time of LFP recording (morning vs evening) had an impact on SDC efficacy. The evening-calculated SDC correlated strongly with the default (morning) SDC marker, (Fig. 4.c.) and predicted the HDRS-17 derived states with an accuracy comparable to the morning-SDC (ACC: 0.74 ± 0.18, n = 7 participants; two-sided Wilcoxon signed-rank test, W = 2.0, *p* = 0.75, r = 0.12, 95% CI = [0.000, 0.039]) (Fig. 4.d., Supp. Fig. 9.). These findings suggest that the SDC remains an accurate indicator of depression state as long as the recording conditions remain consistent. This flexibility in obtaining the SDC offers additional convenience for patients, clinicians and device manufacturers.

### Narrowband spectral power biomarker surrogates

While the previous sections have demonstrated that the SDC is an effective biomarker for tracking core depression state relevant for decision support in SCC DBS, it relies on complex signal features that preclude ongoing tracking given the constraints of current commercially available DBS devices. In particular, the Medtronic Percept™ allows tracking of narrowband power (once every 10 minutes) but not complex combinations of spectral features. It would aid scalability of SCC DBS if narrowband surrogate biomarkers can approximate the SDC in decision support systems.

To investigate this possibility, we performed an exhaustive data-driven search for putative narrowband power surrogate biomarkers of the full SDC by applying a sliding window of 5 Hz bandwidth across the 0–40 Hz frequency range in both hemispheres to calculate the power within each window (detailed in the methods section). The average performance (area under the receiving operating characteristic, AUROC) across the combined cohorts was evaluated for each center frequency to identify the power band that best approximated the SDC by having the best combination of high mean-AUROC and least variance (Fig. 5.a.). The analysis identified left 12–17 Hz and right 10–15 Hz band powers as the best narrowband range in each hemisphere to serve as a putative SDC surrogate. The recovery states identified from the left and right bands matched the sick and stable response states identified by the SDC, achieving high accuracies (ACC: 0.87 ± 0.17 for the left; ACC: 0.88 ± 0.14 for the right) using thresholds determined from the ROC curve (Fig. 5.b.). When compared to HDRS-17-derived states, the left power band-derived states showed numerically lower accuracy (ACC: 0.69 ± 0.27) than the right power band-derived states (ACC: 0.79 ± 0.18), though this difference was not statistically significant (two-sided Wilcoxon signed-rank test, W = 8.5, *p* = 0.11, r = 0.46, n = 12 pairs, 95% CI = [−0.220, 0.021]) (Fig. 5.b., Supp. Fig. 10.). Despite the higher accuracy at capturing HDRS-17 state transitions with a right hemisphere narrowband surrogate signal, the utility of this information for clinical decision support remains to be investigated due to the nonspecific nature of the HDRS-17 and SDC-HDRS-17 incongruence described above (and shown in Fig. 2.c.).

**Figure 5:**
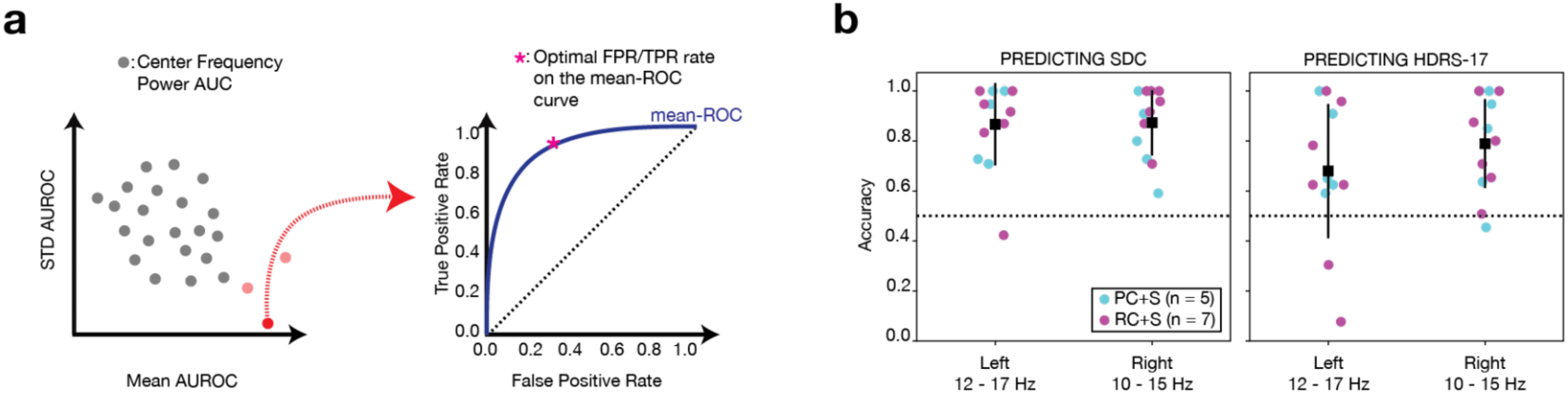
a) Illustration of choosing the center frequency with highest correlation to SDC in each hemisphere and identifying the best threshold for all the participants in both cohorts. b) Performance summary of Left 12-17 Hz and Right 10-15 Hz in predicting the SDC-identified and HDRS-17-identified stable response transitions. Dots indicate individual participants, and color indicates the cohort (Cyan: PC+S, Magenta: RC+S).

## DISCUSSION

In this study, we developed a novel multimodal data collection application for acquiring self-initiated intracranial local field potential recordings, video diaries, and behavioral ratings from 10 new individuals undergoing SCC DBS for TRD. The platform, built on the prototype Medtronic Summit RC+S system, demonstrated that it is possible to collect multimodal data with high compliance rates in SCC DBS participants as well as track LFP changes in a variety of different conditions (stimulation turned ON and OFF, during morning and evening). With no exposure to data from the current cohort for retraining or recalibration, the SDC biomarker derived from the LFP on previous cohorts predicted transitions to stable response with high accuracy, demonstrating the critical generalization of this biomarker from previous findings (Alagapan et al. 2023) necessary for practical utility in a decision support system. Notably, this cohort differed from the prior cohort the SDC was trained on in a variety of ways, including geographical location from which participants were recruited (New York, NY, for the RC+S cohort vs Atlanta, GA, for the PC+S cohort), the DBS device itself (Summit RC+S vs Activa PC+S) with different data acquisition electronics, and the clinical management team (site specific neurosurgeons and psychiatrists). Taken together, this study supports the notion that continuous SCC DBS using connectomic-based targeting (Riva-Posse et al. 2013, 2018; Alagapan et al. 2023) can induce a robust and sustained antidepressant response measurable with a common biomarker collected at home, using only occasional stimulation dose adjustments by the clinical team.

The results from these imaging analyses are also consistent with our previously reported findings (Alagapan et al. 2023), with this expanded analysis strengthening the association between white matter (WM) integrity and the trajectory of recovery mediated by SCC DBS. Specifically, we identified abnormalities in the cingulum bundle, uncinate fasciculus/ventral amygdalofugal pathway, and forceps minor tracts that correlated with the time to stable response in both the PC+S and RC+S cohorts as well as with deficits in functional connectivity between SCC and regions that are connected to the SCC by the respective WM tracts. These findings support distributed dysfunction of a putative limbic-cortical depression network (Mayberg 1997, 2009) anchored by newly identified WM scan abnormalities in strategic locations across specific bundles, consistent with postmortem reports demonstrating decreases in oligodendroglia in ventromedial frontal cortex, subcallosal cingulate and the amygdala in samples with documented antemortem depression (Öngür et al. 1998; Rajkowska et al. 2015).

Taken together, this evidence of disease pathophysiology points to an important hypothesis for future testing: SCC DBS may serve to address this network dysfunction by promoting remyelination across brain networks (including the cingulum bundle), consistent with previous reports in animal models (Fujimoto et al. 2026; Gibson et al. 2014). Robust FA changes accompany natural recovery from injury in humans (Thengone et al. 2016), and SCC DBS selectively increased cingulum FA and oligodendrocyte myelination (Fujimoto et al. 2026) in a nonhuman primate model, aligning with the cingulum localization reported here. The prolonged carryover we observe may reflect such remodeling if the cingulum and forceps minor are under sustained SCC activation, potentially via an interhemispheric SCC-to-SCC connection.

This study provided several opportunities to verify the specificity of the SDC as a depression state tracker, which is an important characteristic for future utility in a decision support system for SCC DBS clinical management. First, as reported previously in a single participant (Extended Data 7 in (Alagapan et al. 2023), we found a subgroup of responders where the week of transition to stable response identified by SDC preceded the transition week identified from HDRS-17 in responding participants. This incongruence corresponded to the recognition of residual anxiety symptoms concurrently with improvement of core depression symptoms (indicated by persistent non-zero scores on the psychic anxiety item in the HDRS-17), similar to observations in earlier SCC DBS cohorts (Crowell et al. 2015).

While origins of the observed anxiety in this cohort are varied, DBS was ineffective in attenuating anxiety symptoms in all cases, as predicted by the stable SDC. Second, several of the case studies also demonstrated clinical instability without anxiety but where the SDC also remained stable, thereby correctly predicting that stimulation adjustments were unnecessary. Retrospective review of the clinical decisions made by the study psychiatrist were also varied, with participant R005 responding to supportive psychotherapy while participant R006 required medication. These case studies demonstrate the utility of a selective biomarker in potentially avoiding needless periods of fruitless stimulation adjustment and delay in appropriate adjunctive treatment.

This cohort also provided several opportunities to verify the sensitivity and stability of the SDC, which is important for understanding the time course of recovery and the timing of potential stimulation adjustments when required. First, the SDC remained stable during a planned one-week stimulation discontinuation after chronic therapy in all subjects. While a one week discontinuation cannot rule out the potential for SDC instability with a longer discontinuation period, the fact that the SDC remained stable even in the minority of patients experiencing brief symptom recurrence (potentially due to a nocebo effect arising from the expectation of discontinuation) is evidence supporting the utility of the SDC at distinguishing relapse requiring intervention from transient distress during stable recovery. Notably, this stability with discontinuation after 6 months of stimulation may be specific to SCC DBS, with other depression DBS targets such as the ventral striatum/ventral capsule appearing to show rapid symptom deterioration in the absence of stimulation (Bergfeld et al. 2016; Wang et al. 2025). Second, one subject showed SDC worsening after a more prolonged unplanned double-blind discontinuation of stimulation 9 weeks post implantation, followed by SDC improvement after stimulation resumption three weeks later.

Interestingly, the SDC had only recently stabilized before stimulation was inadvertently discontinued, suggesting that an emerging SDC can be lost with insufficient stimulation early in therapy but quickly recaptured. This raises the possibility of an SDC-based alarm indicating the need for stimulation adjustments over the course of chronic DBS treatment. Taken together, the specificity, sensitivity and stability demonstrated by the SDC in these analyses support its value for monitoring DBS sensitive and DBS-insensitive brain state changes suitable for a decision support system to optimize comprehensive DBS clinical care.

The biomarker analysis enabled by this in-home data collection system provided two important validations of robustness that will be critical for practical utility. First, while recent studies have highlighted circadian related changes in LFP across SCC (van Rheede et al. 2024), subthalamic nucleus (van Rheede et al. 2022), ventral striatum (Provenza et al. 2024), and impedance (Mivalt et al. 2023), the SDC calculated by normalizing to the respective (morning or evening) baseline recordings showed no significant performance differences with recordings made in the morning or evening. Second, we did not find any significant difference in the predictive performance of SDC derived from recordings made with stimulation on or with stimulation off. These findings are consistent with our observation that SCC LFP exhibits stimulation-induced differences only acutely or in the early stages of therapeutic stimulation (Fitoz et al. 2023). Taken together, these findings suggest the biomarker is robust to the conditions under which the recordings were made and highlight the potential of the SDC to be implemented onboard DBS systems that sample throughout the day without interrupting stimulation.

To move toward scalable decision support systems that can be implemented on commercially available devices such as the Percept™ (Bronte-Stewart et al. 2024), we identified narrowband power changes in each hemisphere that are the best available surrogate biomarkers for the SDC. The beta and alpha bands identified in this analysis have been previously found to be responsive to acute stimulation (Fitoz et al. 2023; Sendi et al. 2021; Smart et al. 2018), and left hemisphere beta band changes in particular were a major contributor to SDC in the PC+S cohort (Alagapan et al. 2023). Beyond SCC, these bands have been associated with mood symptoms across humans and animal models (Xiao et al. 2023; Hultman et al. 2018; Benschop et al. 2021; Kirkby et al. 2018; Xiao et al. 2024). While both potential surrogates captured SDC transition times with high accuracy, right alpha captured more of the fluctuations in the HDRS-17 transition times than left beta. This preliminary observation coupled with the symptom specificity of the SDC shown in Fig. 2.c. raises the possibility of discovering additional biomarkers that capture co-occurring distress or other non-specific depression symptoms (e.g., anxiety, rumination, etc.). In an exploratory analysis, we correlated baseline normalized power across the full 0 to 40 Hz frequency range with the baseline normalized psychic anxiety subitem of the HDRS-17 over the observation period, contrasting the responders exhibiting persistent anxiety against the remaining participants. Correlations in the lower frequencies were numerically more positive in the anxiety persistent subgroup, but no center frequency reached significance in a between group comparison (all p > 0.05, two sided Mann Whitney U test). This null result, together with the heterogeneous and partly pharmacological nature of the symptoms observed in participants R005 and R006, suggests that capturing these DBS insensitive symptoms may require markers beyond the SCC LFP features used here, potentially including dedicated symptom instruments or multimodal behavioral features such as language and facial expression derived from the video diaries.

Nonetheless, this hypothesis is consistent with other observations of right hemisphere electrophysiology being broadly associated with emotional reactivity (Scangos et al. 2021), anxiety (Kirkby et al. 2018; Wang et al. 2025), OCD (Provenza et al. 2021, 2024), and trauma symptoms (Gill et al. 2023). In particular, the observation that theta band activity in BNST DBS subjects (Wang et al. 2025) tracked with momentary anxiety changes provides motivation for ongoing research to identify biomarkers of transient distress that are distinct from biomarkers of core depression state. While this preliminary analysis supports the continued investigation of individual power bands in the SCC, significant further investigation is necessary to establish confidence in single band signals to inform a decision support system.

The current study has several limitations. First, the analyses presented here were retrospective, with all clinical decisions regarding DBS dose or medications adjustments made by the study psychiatrist without access to the LFP biomarkers. Future studies will require prospective testing to evaluate SDC using a well-defined decision support system during the various clinical scenarios arising during ongoing DBS therapy. Finally, while the limited sample size of 7 individuals in the RC+S cohort is relatively small to confirm the generalizability of the SDC to a broader population, the performance of the models (which have never been exposed to the data from the RC+S) supports the generalizability of the core biomarker findings. As both the PC and the RC platforms are retired, these findings do provide reasonable incentive to retain the model and test both the SDC and the surrogate biomarkers in the commercial Percept™ system before widespread use.

## Data Availability

The data that support the findings of this study will be publicly available via the Data Archive for The Brain Initiative (DABI).

## ACKNOWLEDGEMENTS

The authors thank Fawad Jamshed and Muhammad Furqan Afzal for assistance in the Mount Sinai Summit Home Application programming, as well as Scott Stanslaski (Medtronic, Inc.), Ben Isaacson (Medtronic, Inc.) and the OpenMind consortium (supported by NIH Brain Initiative through NINDS grant U24NS113637) for technical support provided during the app development and collection of LFP data. The authors also acknowledge the technical and platform support of Rune Labs for LFP data management. The authors acknowledge the expertise of Lisa Feldman Barrett in designing the visual analog scales for the EMA. This work was supported by the NIH BRAIN Initiative through NINDS grant numbers UH3NS103550 and UH3NS141080, the NIH National Center for Advancing Translational Sciences grant numbers UL1TR002378 and KL2TR002381, the Hope for Depression Research Foundation, the Wellcome Leap Multi-Channel Psych (MCPsych) Program, James S. McDonnell Foundation grant number 220020399, National Science Foundation CAREER award number CCF-1350954, and the Julian T. Hightower Chair at Georgia Tech. The Activa PC+S devices and Summit RC+S devices were provided by Medtronic, Inc. in coordination with the UH3 grant funding. The content is solely the responsibility of the authors and does not represent the official views of the National Institutes of Health (NIH) or Medtronic, Inc.

## AUTHOR CONTRIBUTIONS

HM, CR, SA, PRP, MF, KC, SH, AW designed the study. MO, TD, SA developed the MSSHA app.

MO, SH, TN, RG, AR, MK, GF collected LFP data. JC, KC acquired neuroimaging data. MF, BK, IT, JG, JA, SN, AW collected clinical assessments. SH, AR, IT curated the data.

ECF, SA, CR, HM, PS designed, performed and evaluated analysis of LFP data. JC, KC, HM, HS designed and performed analysis of neuroimaging data. ECF, SA, CR, HM, MF, PRP designed and performed analysis of clinical data.

ECF, SA, CR, HM, JC, KC prepared the manuscript.

All authors provided substantive feedback for revision of manuscript.

## METHODS

### Participants and Clinical Assessments

Ten consecutive participants with treatment-resistant major depressive disorder who met eligibility criteria were enrolled in a single-group, open-label trial involving a single active DBS intervention that enabled local field potential collection from the stimulation site (ClinicalTrials.gov identifier NCT04106466). The eligibility criteria and sample size were selected in accordance with previously established work (Holtzheimer et al. 2012; Riva-Posse et al. 2018; Alagapan et al. 2023). Written informed consent was obtained from all participants. The protocol was approved by the Institutional Review Board at the Icahn School of Medicine at Mount Sinai, and the US Food and Drug Administration under a physician-sponsored Investigational Device Exemption (IDE G130107) and was monitored by the Emory University Department of Psychiatry and Behavioral Sciences Data and Safety Monitoring Board.

Clinical symptom severity was evaluated weekly using the 17-item Hamilton Depression Rating Scale (HDRS-17), the Montgomery-Åsberg Depression Rating Scale (MADRS), and the Beck Depression Inventory (BDI). Additional daily assessments included the Computerized Adaptive Testing for Mental Health (CAT-MH; Adaptive Testing Technologies, Chicago, Illinois) and Visual Analog Scale (VAS) of mood, motivation, energy, and alertness. Participants were interviewed weekly to monitor HDRS-17 changes and make stimulation adjustments as needed. All clinical decisions (including DBS, medication or psychotherapy changes) were made by the study psychiatrist and were not automatic adjustments based on the LFP biomarker.

The definitions of "response" and "remission" follow those outlined previously (Alagapan et al. 2023). Clinical and demographic data for the participants are listed in Table 1. Individually, of the ten participants, eight responded to the therapy (more than 50% decrease from the presurgical baseline in HDRS-17), and four met remission criteria (HDRS-17 less than 8). Three participants were excluded from the analysis due to significant shifts in canonical frequency band powers within the LFP data, which are essential for the analysis. These abrupt signal changes were not attributable to any identifiable clinical or behavioral events and the precise mechanism remains uncertain. A custom moving window-based shift detection algorithm was employed to identify shifts in dB scale power persisting for a week or longer. A shift was identified as a 3 dB increase/decrease in the power of the current day from the 7-day window average (Supp. Fig. 4. and Fig. 1.d.). We also examined lead impedance around the times of these shifts.

**Table 1:** Patient Demographics.

| Participant ID | Age at time of Surgery (years) | Sex | HDRS-17 (Pre) | HDRS-17 (End) | Stim Start | Stim End | Number of Stim Changes | Duration of Current Episode (months) | Age at first depressive episode (years) | Number of Depressive Episodes | Reason for Exclusion | Responder Status |
| --- | --- | --- | --- | --- | --- | --- | --- | --- | --- | --- | --- | --- |
| R001 | 31-35 | F | 28 | 16 | 4.5 mA | 8 mA | 1 | 108 | 16-20 | 2 | N/A | Nonresponder |
| R002 | 51-55 | F | 22.75 | 12 | 4.5 mA | 5 mA | 1 | 144 | 21-25 | 2 | N/A | Responder* |
| R003 | 26-30 | M | 24.5 | 11 | 4.5 mA | 4.5 mA | 0 | 48 | 11-15 | 4 | N/A | Responder |
| R004 | 61-65 | M | 28 | 9 | 4.5 mA | 4.5 mA | 0 | 60 | 11-15 | N/A | Right Hem Rec. Artifact | Responder |
| R005 | 36-40 | F | 23 | 5 | 4.5 mA | 4.5 mA | 0 | 24 | 16-20 | 5 | N/A | Responder |
| R006 | 41-45 | M | 21.5 | 7 | 4.5 mA | 7 mA | 3 (2I, 1D) | 108 | 21-25 | 4 | N/A | Responder |
| R007 | 31-35 | M | 23.5 | 10 | 4.5 mA | 4.5 mA | 0 | 72 | 6-10 | 2 | Impedance Drop Artifact | Responder |
| R008 | 46-50 | M | 23 | 6 | 4.5 mA | 4.5 mA | 0 | 122 | 16-20 | 3 | Both Hem Rec. Artifact | Responder |
| R009 | 36-40 | M | 23.5 | 18 | 4.5 mA | 6 mA | 1 | 12 | 11-15 | 4 | N/A | Nonresponder |
| R010 | 36-40 | F | 21.5 | 2 | 4.5 mA | 4.5 mA | 0 | 24 | 11-15 | 4 | N/A | Responder |

\* Participant R002 met the response criterion at week 25 (one week after the predefined endpoint and before the discontinuation phase) and was included as a responder in all analyses.

Although impedance changes were observed, they were neither concurrent with nor consistent in direction with the spectral power shifts, preventing clear attribution of these power changes to impedance changes. All the three excluded participants were classified as responders based on HDRS-17 scores (Supp. Fig. 5.).

Among the seven participants with usable electrophysiology data, four had a discontinuation period extending beyond six months due to their response to DBS therapy, with discontinuation occurring at week 37, week 26, week 25 and week 25, respectively. The remaining participants entered the discontinuation period immediately following six months of chronic stimulation (week 24).

### Summit RC+S Application Development

The Medtronic Summit RC+S system is a second-generation bidirectional DBS system that allows streaming data collection from the contacts of the implanted DBS lead. The system can be controlled using an application programming interface (API) that is accessed using a ‘research development kit (RDK; Medtronic model 4NR013)’. The RDK allows the configuration of stimulation and sensing as well as initiating both functions. We leveraged this capability to develop applications that supported the data collection for this study. The Mount Sinai depression DBS platform consists of two main applications: home-based and research application. The home-based application is installed on a Windows support tablet that participants can take home, while the research application is used within the laboratory. The process of recording LFP data is initiated with the decision tree of whether the participant is visiting the laboratory for an experiment session. If the participant is visiting the laboratory for an experiment, the stimulation parameters will need to be adjusted first using the Research Lab Programmer (RLP). The clinician sets the parameter bounds in which a technician can safely deliver stimulation. When the participant is not visiting the laboratory, LFP data will be recorded using the home-based application.

Both the home-based and research applications were developed based on the Agile development methodology. User and system requirements were captured during the design and analysis phase. Careful risk analysis was done, risk-benefit analysis was performed, and sets were drawn to mitigate risk analysis with the support of medically trained clinicians. At the end of each prototype, verification was performed using unit testing and integration testing. At the end of the prototype phase, validation was performed of the system requirements. Guidance from the regulatory quality management system was taken to increase security and participant safety and to minimize the threat of malicious hacks, system malfunctioning, and failure. Some of the main regulations used were IEC 62304 Medical Device Software Development, ISO 13485 - Medical Device Development, and ISO 14971 - Medical Devices Risk Management Assessment. (ISO-13485, ISO-14971, ISO 13485).

### SCC DBS and LFP recording

The surgical protocol for electrode implantation followed previously published work (Alagapan et al. 2023; Riva-Posse et al. 2018).Medtronic 3387 leads were implanted in subcallosal cingulate cortex targets informed by diffusion MRI at the intersection of three critical white matter pathways: the forceps minor, cingulum bundle, and uncinate fasciculus/ventral amygdalofugal pathways (midline frontostriatal fibers). The comparative analysis of the target locations, as observed through imaging, is provided in Fig. 1.a., Supp. Fig. 1. and Extended Table 2.. Stimulation was delivered through the Summit RC+S system, using a current-controlled pulse generator. The stimulation frequency was set to 130 Hz with a pulse width of 90 µs for all participants and was not changed during therapy. The initial stimulation amplitude was set to 4.5 mA with subsequent changes determined by the study psychiatrist. Details of the stimulation amplitude changes are provided in Table 1. Chronic therapeutic stimulation in the first 5 participants was initiated 30 days postsurgery, following previously published studies (Alagapan et al. 2023; Riva-Posse et al. 2018). This postsurgery recovery phase was discontinued for the second 5 participants and chronic therapeutic stimulation was initiated immediately following surgery.

Each participant was provided with a tablet containing a patient-specific application for the collection of neural and behavioral data at home. This application is designed to facilitate recording sessions twice daily, specifically during morning and evening periods throughout the observation period. In addition to these regular recording sessions, the home-based system incorporates daily video recordings to complement the electrophysiological data collection. Overall experimental setup scheme is presented in Fig. 1.b.. Recording sessions lasted approximately eight minutes, and differential LFPs were collected from the side contacts adjacent to the stimulation contacts. The LFP recordings consisted of three distinct phases: a three-minute stimulation-on period, followed by a three-minute stimulation-off period, and concluding with two minutes of stimulation-on post-off. The recorded data is synced over the cloud for storage.

### Feature Extraction and Biomarker Estimation

Raw time-domain LFP signals are extracted from the recorded data using the open-source toolbox used for RC+S data analysis (Sellers et al. 2021). The LFP obtained during both the stimulation-on and stimulation-off periods were segmented into 10-second intervals. In line with the methodology used previously (Alagapan et al. 2023), only the stimulation-off periods were considered for the primary analysis. Recordings from the stimulation-on period preceding the stimulation-off period were processed separately and analyzed for comparison with the stimulation-off results. The first and last segments in both conditions (10 seconds) were discarded to account for switching-related artifacts.

Features were extracted from the LFP to estimate the previously identified Spectral Discriminative Component (SDC) biomarker. The SDC is a low-dimensional representation of the changes in LFP features accompanying stable recovery and estimated using a Generative Causal Explainer (GCE) framework detailed previously (Alagapan et al. 2023; O’Shaughnessy et al. 2020). Higher SDC values indicate the ’sick’ state and lower values indicate the ’stable response’ state. The 10-second LFP segments were processed using Nitimes multi-taper fast Fourier transform with adaptive weights to obtain features for the SDC. The feature set comprised spectral power and coherence at canonical frequency bands for both hemispheres (delta: 1–4 Hz; theta: 4–8 Hz; alpha: 8–13 Hz; low beta: 13–20 Hz; high beta: 20–30 Hz; gamma: 30–40 Hz), and phase-amplitude coupling between the delta band (1.5–3.0 Hz) and the high beta/gamma band (20–35 Hz), calculated using PACtools. This process yielded a 20-dimensional feature set.

The features were baseline-corrected relative to the first week of chronic stimulation (C1) rather than the postsurgery baseline (B4) used in previous analyses. This adjustment ensures consistent preprocessing for the RC+S cohort, as the last five participants began chronic stimulation immediately after surgery. To enable a comparable measure for the PC+S cohort, the baseline was similarly shifted from B4 to C1, and the SDC was recalculated. Performance with C1 normalization in the PC+S cohort remained comparable (AUC 0.98 ± 0.008 for C1 vs. AUC 0.94 ± 0.04 for B4; two-sided Wilcoxon signed-rank test, W = 1.0, *p* = 0.1250, r = 0.69, n = 5 pairs, 95% CI = [−0.021, 0.085]) and yielded identical transition weeks. Therefore, C1 normalization is adopted in both PC+S and RC+S cohorts for subsequent analyses without significant performance changes.

Consistent with prior work (Alagapan et al. 2023), each feature underwent min-max scaling over the initial 24-week period as a preprocessing step. The derived scaling was subsequently applied to features beyond the 24-week period until treatment discontinuation in R001, R002, R009 and R010.

Following this, the scaled features were projected through the feature compression network of the GCE model trained on the PC+S cohort data (Alagapan et al. 2023) to obtain SDC, which is then transformed to indicate the probability of being in the ‘sick’ state. Post-transformation, the obtained SDC values for each segment were averaged across each week to derive weekly SDC values. For stable transition week identification, a threshold of 0.5 was previously applied to the SDC, as it showed comparability with the HDRS-17 measure. This threshold was further validated by binarizing the weekly SDC and the HDRS-17 identified stable response state in the PC+S cohort and selecting the threshold at the optimal operating point of the averaged ROC curve, which confirmed 0.5 as optimal. The same ROC based procedure was used to determine the universal thresholds for the narrowband 5 Hz power surrogates described below.

### Determining single frequency band surrogate of SDC biomarker

Power in 5-Hz bands from the left and right hemisphere LFP that can capture the complex SDC biomarker were determined to enable the tracking of LFP biomarkers in commercially available bidirectional DBS devices. Power spectra computed for the estimation of SDC were used to obtain 5 Hz bands over center frequencies ranging from 2.5 Hz to 37.5 Hz. These power values were subsequently baseline-corrected with respect to the first week of the chronic stimulation period to obtain z-scores. The z-scores were then scaled over the 24-week period. The transition week for power was estimated to occur when the power exceeded the established threshold and did not drop below this threshold for more than two consecutive weeks. This approach contrasts with the approach previously used for determining SDC and HDRS-17 transition weeks (Alagapan et al. 2023) in which the transition week was determined as the week where the values fell below the threshold and remained below the threshold. This approach was motivated by the observation that increases in power accompanied stable recovery in contrast to SDC and HDRS-17 where decreases accompanied stable recovery.

To determine optimal thresholds for transition week calculations based on weekly power values, a comprehensive threshold sweep was conducted from 0 to 1 with 0.1 increments. Utilizing the identified transition weeks, the observation period was binarized into two states: ‘sick’ until the identified transition, and ’well’ thereafter. These binarized state vectors were subsequently compared against the states predicted by SDC, which served as the ground truth.

For each subject, Receiver Operating Characteristic (ROC) curves were computed across the tested thresholds, revealing optimal individual thresholds that maximized performance in estimating transition weeks based on power bands. This subject-specific analysis provided insight into individual variability in electrophysiological responses to treatment. To establish a universally applicable threshold across the combined PC+S and RC+S cohorts, mean Area Under the Curve (AUC) values were analyzed. This analysis yields a universal threshold optimized for all subjects at specific center frequencies. For each individual power band, the mean AUC with the optimal universal threshold and its standard deviation across subjects were recorded, providing a measure of the consistency of each power band as a predictor of SDC state transitions. Finally, the 5-Hz band was chosen among the three center frequencies with the highest mean AUCs that exhibited the lowest deviation across participants. This approach ensured that the selected center frequency and threshold are both unbiased and consistent across the entire cohort. This process resulted in identification of two center frequencies capable of tracking SDC-defined transitions across hemispheres: 14.5 Hz in the left hemisphere (AUC: 0.89 ± 0.17) and 12.5 Hz in the right hemisphere (AUC: 0.95 ± 0.03), with respective thresholds of 0.4 and 0.5.

### Comparison against Clinical Assessment Measures

The SDC for the RC+S cohort was compared against the HDRS-17-based stable response week to evaluate the efficacy of the biomarker in this cohort. For HDRS-17 comparisons, we established a normalized baseline using the average pre-surgical HDRS-17 scores. Stable clinical response was defined as a sustained reduction in the relative HDRS-17 score below 0.5 that persisted without exceeding this threshold for more than two consecutive weeks.

Similarly, for CAT-MH comparisons, we utilized the CAT-DI depression severity percentage measure, scaling scores from their original 0-100 range to 0-1, and calculated weekly averages. The threshold for stable response was set at 0.5, corresponding to the "Mild" symptom severity on the CAT-MH scale. Accuracy of SDC and single-band surrogates in predicting stable response against these clinical measures is reported in Extended Table 1.

For the specificity analysis, we included the responders in whom the SDC identified stable response transition preceded the HDRS-17 identified transition (R002, R003, R005, R006). Said another way, we included all participants in whom there was a period where the SDC and HDRS-17 were incongruent.

### Image Acquisition and Processing

High-resolution structural T1-weighted (T1w), resting-state functional MRI (fMRI), and diffusion-weighted images (DWI) data were acquired using two scanners: a 3T Siemens Prisma MRI scanner (Siemens Medical Solutions) and a 3T GE Signa Architect scanner (GE HealthCare). T1w images were collected using a 3D magnetization-prepared rapid gradient-echo (MPRAGE) sequence with the following parameters (Prisma/Signa): sagittal slice orientation; resolution=1×1×1mm3/0.977×0.977×0.6mm3; repetition time (TR)=2600/8.432ms; inversion time (TI)=900/1100ms; echo time (TE)=3.02/3.188ms; and flip angle=8°. Resting-state fMRI data were acquired with participants’ eyes open using a gradient echo-planar imaging (EPI) sequence with the following parameters (Prisma/Architect): number of volumes=460/150; number of axial slices=56/60; voxel size=2×2×2mm3/2.5×2.5×2.5mm3; TR=1000/2000 ms; and TE=30 ms. DWI data was acquired using a single-shot spin-echo EPI sequence with the following parameters (Siemens Prisma or GE Signa Architect): 60 non-collinear directions with five non-diffusion weighted images (b0); b-value=1000s/mm2 (Prisma) and 1200s/mm2 (Architect); number of slices=66/80; voxel size=2×2×2 mm3; TR=3292/17000ms; TE=96/78.3ms. An additional five non-diffusion weighted image datasets with opposite phase encoding were also collected to compensate for the susceptibility-induced distortion.

All images were preprocessed using the FMRIB Software Library (FSL; http://www.fmrib.ox.ac.uk/fsl/) (Jenkinson et al. 2012) (v6.0) and Analysis of Functional NeuroImages (AFNI, https://afni.nimh.nih.gov) software (v23.1.06). T1w image was skull stripped and normalized to the MNI152 template using the fsl_anat toolbox and ANTs. The standard preprocessing pipeline for fMRI, including despiked and corrected for slice time acquisition differences and head motion, implemented in the AFNI was used for resting-state fMRI preprocessing. The remaining effect of noise signals, including head motion inferences, signals from the CSF, and local white matter, were removed. Subsequently, the data were band-pass filtered (0.01 < f < 0.1 Hz) and spatially smoothed up to 8mm full width at half-maximum (FWHM) using 3dBlurToFWHM in AFNI. The preprocessed fMRI data were normalized to the MNI152 template using previously generated T1w normalization warp-fields. The mean time series of the bilateral SCC seed (±6, +24, -11) (Dunlop et al. 2017) was correlated voxel-wise with the rest of the brain. The voxel-wise correlation coefficient maps were then converted to z-scores by Fisher’s r-to-z transformation. The z-score determined the level of functional connectivity of the SCC seed. DWI data underwent denoise, distortion and motion correction, and bias correction using the MRTrix toolbox and a local tensor fitting (fdt, FSL) to calculate the FA map. Tract-Based Spatial Statistics (TBSS) processing was performed for group analysis (Smith et al. 2006). Briefly, individual FA images were aligned to the standard FMRIB58 FA template using a nonlinear registration, and the normalized FA images were then averaged to create a mean FA image. The mean FA image was thinned to create a FA skeleton representing WM tracts common to all patients. A threshold value of 0.2 was used to exclude adjacent gray matter or cerebrospinal fluid voxels. A volume of tissue activated (VTA) was generated using the Lead-DBS software (Noecker et al. 2018) with patients’ specific chronic stimulation settings (i.e., 130 Hz, 3.5 V, 90 µs). White matter tracts passing through VTA were extracted in each participant using the Xtract toolbox in FSL (Warrington et al. 2020) and then averaged to generate a white matter tract mask that represents the common activation pathways of all five participants. Three white matter masks, including forceps minor (FM), cingulum bundle (CB), and uncinate fasciculus/ventral amygdalofugal pathway (UF/vAFP), were included for the statistical analysis.

Within the specific tracks of the FA skeleton, Spearman’s rank correlation between white matter integrity measure (FA) and the time to stable response, as defined by the SDC, were performed to investigate the relationship between time to stable response and white matter integrity. In parallel, voxel-wise Spearman’s rank correlation analyses were conducted between time to stable response and SCC functional connectivity (FC) to identify functional connections associated with response dynamics. To assess structure–function relationships, Spearman’s rank correlations were further performed between FA values and SCC FC in spatially adjacent brain regions that demonstrated significant associations with time to stable response in the prior analyses.

### Statistical Analysis

All statistical comparisons used non-parametric tests to account for the small sample sizes across both cohorts (n = 5 to 7), which preclude reliable assessment of distributional assumptions. Two-sided tests were used throughout with a significance threshold of α = 0.05. Given the limited power to detect small effects at these sample sizes, descriptive statistics including accuracy and 95% bootstrap confidence intervals are also reported to support the interpretation of comparability across conditions.

To test whether the PC+S and RC+S cohorts followed similar clinical courses, we compared four summary measures between cohorts using two sided Mann Whitney U tests: baseline severity (mean Phase A HDRS-17), endpoint severity (HDRS-17 at week 24), time to stable response, and the Illness Density Index (IDI), the baseline normalized, time averaged HDRS-17 over the observation period (Kelley et al. 2012). The cohorts did not differ significantly on any measure (baseline, *p* = 0.25; endpoint, *p* = 0.26; time to stable response, *p* = 0.17; IDI, *p* = 0.62). Weekly trajectories were additionally modeled with a linear mixed effects model of HDRS-17 including fixed effects of week, cohort, and their interaction, and a random intercept per participant (HDRS-17 ∼ week × group + (1 | subject)).

Comparison of SDC-derived prediction accuracy against HDRS-17 between cohorts was performed using a Mann–Whitney U test (PC+S: n = 5, RC+S: n = 7). Biomarker robustness was assessed through paired Wilcoxon signed-rank tests, comparing the prediction performance of the default SDC against the SDC calculated from stimulation-on, morning recordings and from stimulation-off, evening recordings, to evaluate sensitivity to stimulation state and time of day, respectively. The stimulation state comparison included the six analyzable participants with consistent stimulation-on recordings across the observation period (RC+S, n = 6 pairs); participant R001 was excluded because a stimulation contact change prevented consistent stimulation-on recordings. The time of day comparison retained all seven analyzable participants (RC+S, n = 7 pairs). The effect of feature normalization baseline on SDC performance in the PC+S cohort was tested by comparing AUC-ROC scores when shifting the baseline from B4 to C1 (PC+S, n = 5 pairs). For all tests, the test statistic, exact p-value and effect size are reported.

