## Supplementary Material for "Common Electrophysiology Biomarkers Collected at Home Robustly Track Depression Recovery With Deep Brain Stimulation"

**Extended Table 1. Accuracy Results of Comparisons:**

| **Performance** | **PC+S** | **RC+S** | **TOTAL** |
| --- | --- | --- | --- |
| SDC (STIM OFF, Morning) vs HDRS-17 | 0.85 ± 0.106 | 0.72 ± 0.161 | 0.79 ± 0.156 |
| SDC (STIM OFF, Evening) vs HDRS-17 | - | 0.74 ± 0.179 | - |
| SDC (STIM ON, Morning) vs HDRS-17 | - | 0.83 ± 0.094 | - |
| Left Power vs HDRS-17 | 0.77 ± 0.157 | 0.62 ± 0.316 | 0.685 ± 0.273 |
| Left Power vs SDC | 0.88 ± 0.127 | 0.86 ± 0.188 | 0.87 ± 0.166 |
| Right Power vs HDRS-17 | 0.78 ± 0.209 | 0.79 ± 0.162 | 0.79 ± 0.183 |
| Right Power vs SDC | 0.81 ± 0.149 | 0.92 ± 0.097 | 0.88 ± 0.135 |
| SDC vs CAT-MH | - | 0.71 ± 0.204 | - |
| Left Power vs CAT-MH | - | 0.65 ± 0.343 | - |
| Right Power vs CAT-MH | - | 0.70 ± 0.217 | - |

**Extended Table 2. Comparison of XYZ coordinates between responders and non-responders**

| **Test** | **Stimulation contact** | | **Lower recording contact** | | **Upper recording contact** | |
| --- | --- | --- | --- | --- | --- | --- |
|  | Left | Right | Left | Right | Left | Right |
| Hotelling's T-squared test | T2(3,18) = 6.60, *p* = 0.16 | T2(3,18) = 2.64, *p* = 0.52 | T2(3,18) = 4.69, *p* = 0.28 | T2(3,18) = 2.13, *p* = 0.60 | T2(3,18) = 5.67, *p* = 0.21 | T2(3,18) = 2.37, *p* = 0.56 |
| Non-parametric comparison in each axis | X: *p* = 0.53  Y: *p* = 0.24  Z: *p* = 0.17 | X: *p* = 0.46  Y: *p* = 0.60  Z: *p* = 0.83 | X: *p* = 0.67  Y: *p* = 0.40  Z: *p* = 0.29 | X: *p* = 0.75  Y: *p* = 0.60  Z: *p* = 0.83 | X: *p* = 0.53  Y: *p* = 0.11  Z: *p* = 0.24 | X: *p* = 0.75  Y: *p* = 0.53  Z: *p* = 0.92 |

The locations of stimulation contacts and recording contacts (upper and lower) were compared between responders (n = 17) and non-responders (n = 3) using Hotelling’s T-squared tests and non-parametric rank-sum tests.

**SUPPLEMENTARY FIGURES**

**Supp. Fig. 1. Electrode Localizations**

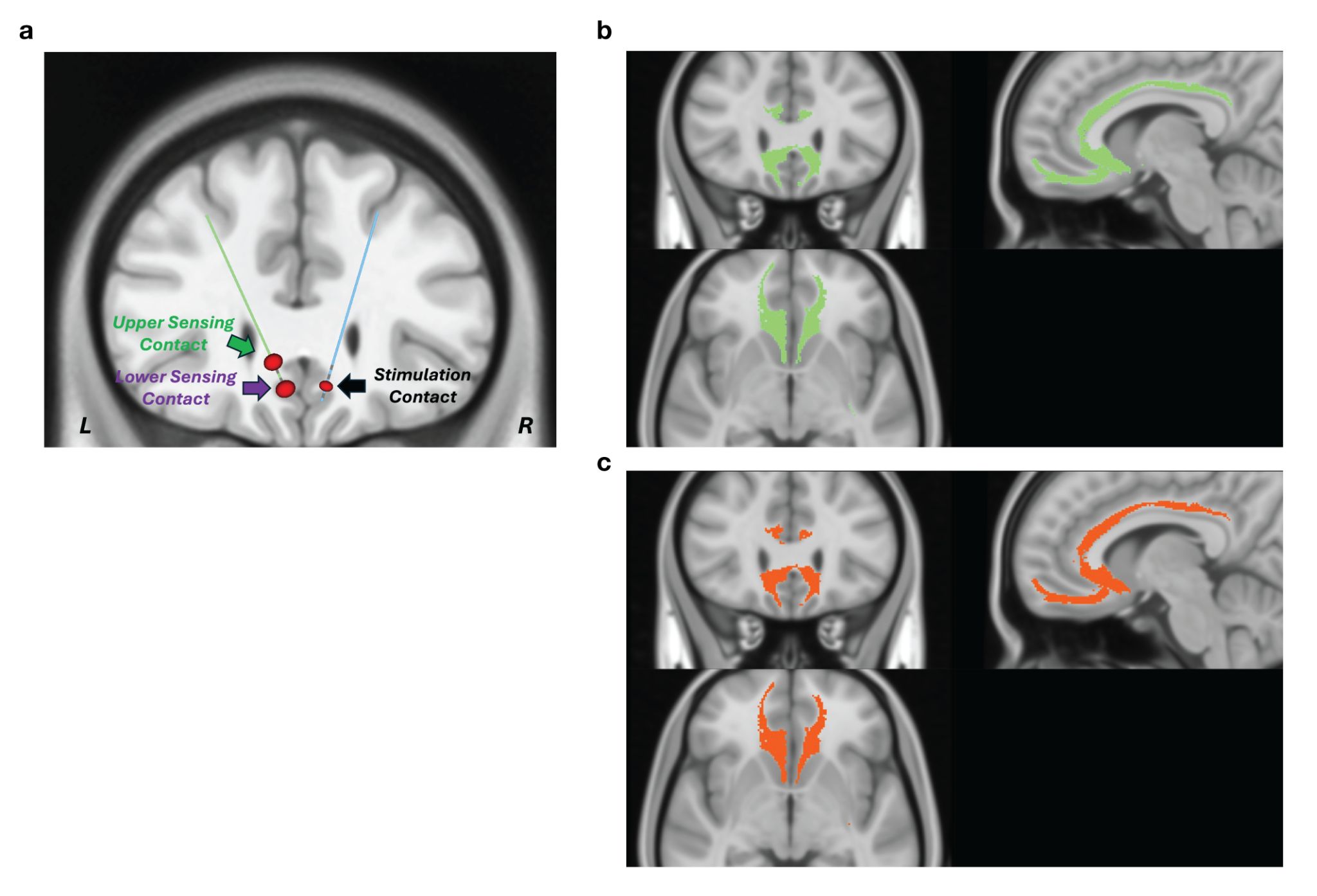

(a) Coronal view depicting reconstructed electrode positions for all subjects in the PC+S and RC+S cohorts. Recording contacts are shown in green (upper sensing contact) and purple (lower sensing contact) and stimulation contact is shown in black. Across subjects, the lower recording contact was consistently positioned within the ventral bank of the subcallosal cingulate cortex. Non-responders demonstrated no deviation from responders in electrode positioning or spatial relationship to the established stimulation field, indicating that treatment outcome differences were not attributable to anatomical targeting variation.
(b) Common voxels stimulated by all responders (n = 17) in both cohorts (PC+S, RC+S), identified using probabilistic connectivity analysis with patient-specific volumes of tissue activated as seed regions.
(c) Common voxels stimulated by all subjects (n = 20) in both cohorts, identified using the same approach as in panel (b). The common voxel map of all subjects showed a high degree of overlap (94.8%) with the responder common voxel map (panel b), indicating that both responders and non-responders exhibited similar white matter stimulation patterns with engagement of the therapeutic white matter pathways. L: left; R: right; A: anterior; P: posterior; S: superior; I: inferior.

**Supp. Fig. 2. Mean HDRS-17 Trajectories**

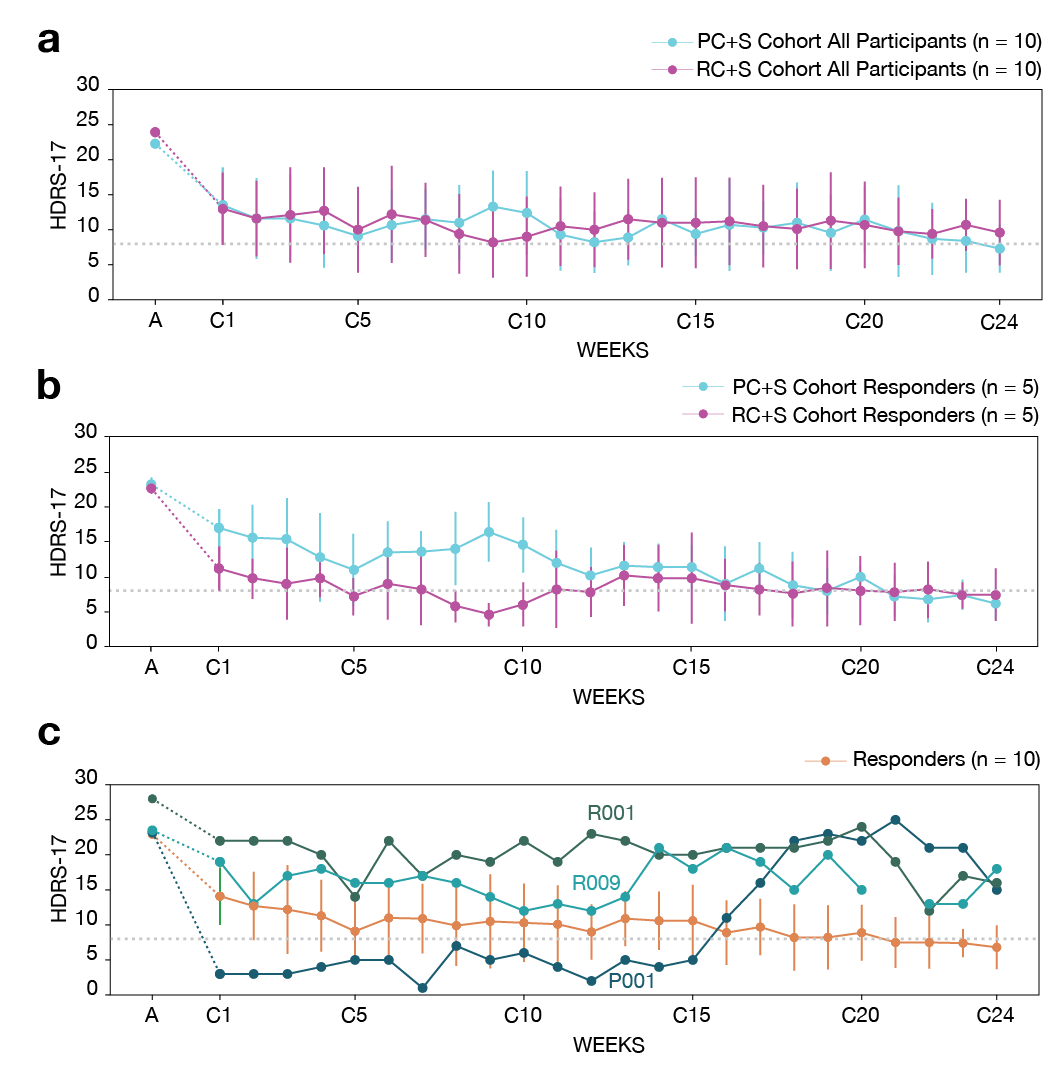

(a) Mean trajectory of HDRS-17 scores over 24 weeks for all participants in both cohorts (PC+S, RC+S).
(b) Mean trajectory of HDRS-17 scores over 24 weeks for responders in the PC+S and RC+S cohorts with usable LFP recordings (included in the analysis).
(c) Mean ± s.d. HDRS-17 trajectory for responders (orange, n = 10) shown with the individual nonresponder trajectories, each in a distinct color. The dotted horizontal line marks the remission threshold (HDRS-17 = 8). The nonresponders follow individually distinct courses with no coordinated change.

**Supp. Fig. 3. Individual and Group Compliance Rates**

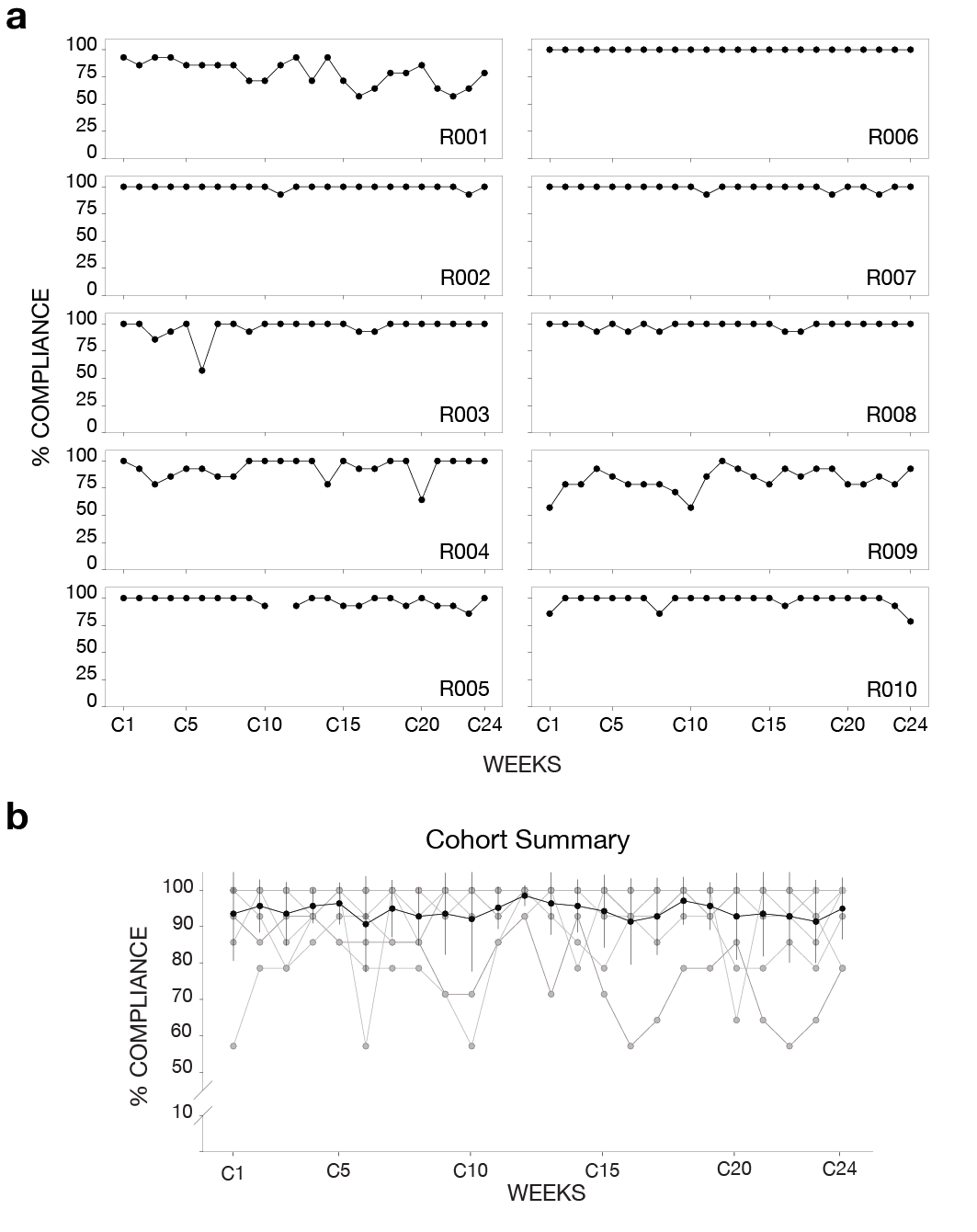

(a) Weekly compliance rates over observation period for each participant.
(b) Aggregated summary, showing above 90% cohort level compliance.

**Supp. Fig. 4. Excluded Participants Spectral Power Shifts**
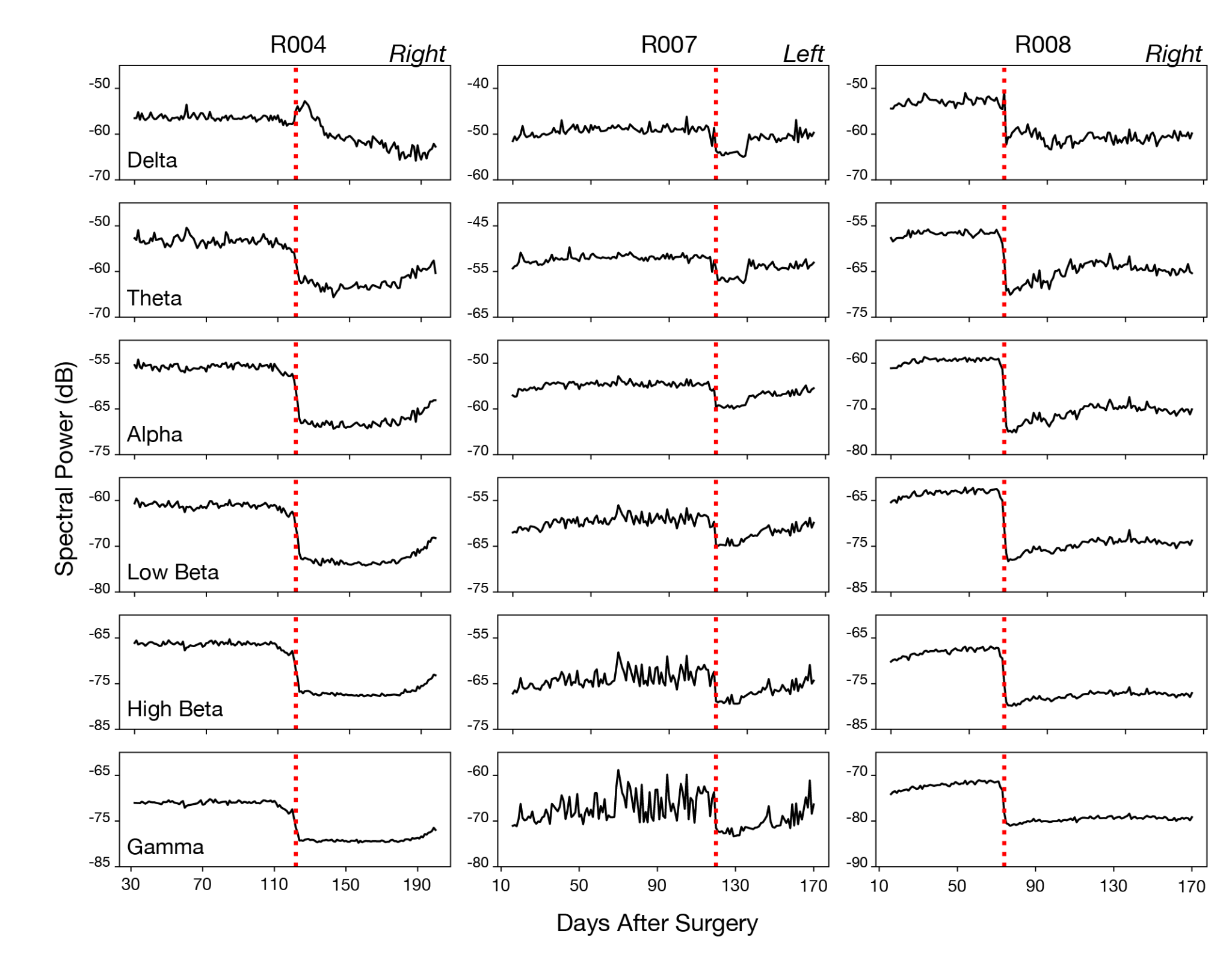

Power shifts detected in participants excluded from analysis, shown for the affected hemisphere in each case (labeled in the corresponding panel). Participant R004 shows power shift more than 3dB for every canonical frequency band in the right hemisphere, showing recordings contaminated on right, not shifting back to its state. R007 shows artifact related power shifts in canonical bands on the left hemisphere; the ongoing effect continues for the rest of the analysis period. Similarly, R008 shows the same characteristics of broadband power shift in the right hemisphere, affecting signal quality.

**Supp. Fig. 5. Excluded Participants HDRS-17 Trajectory**

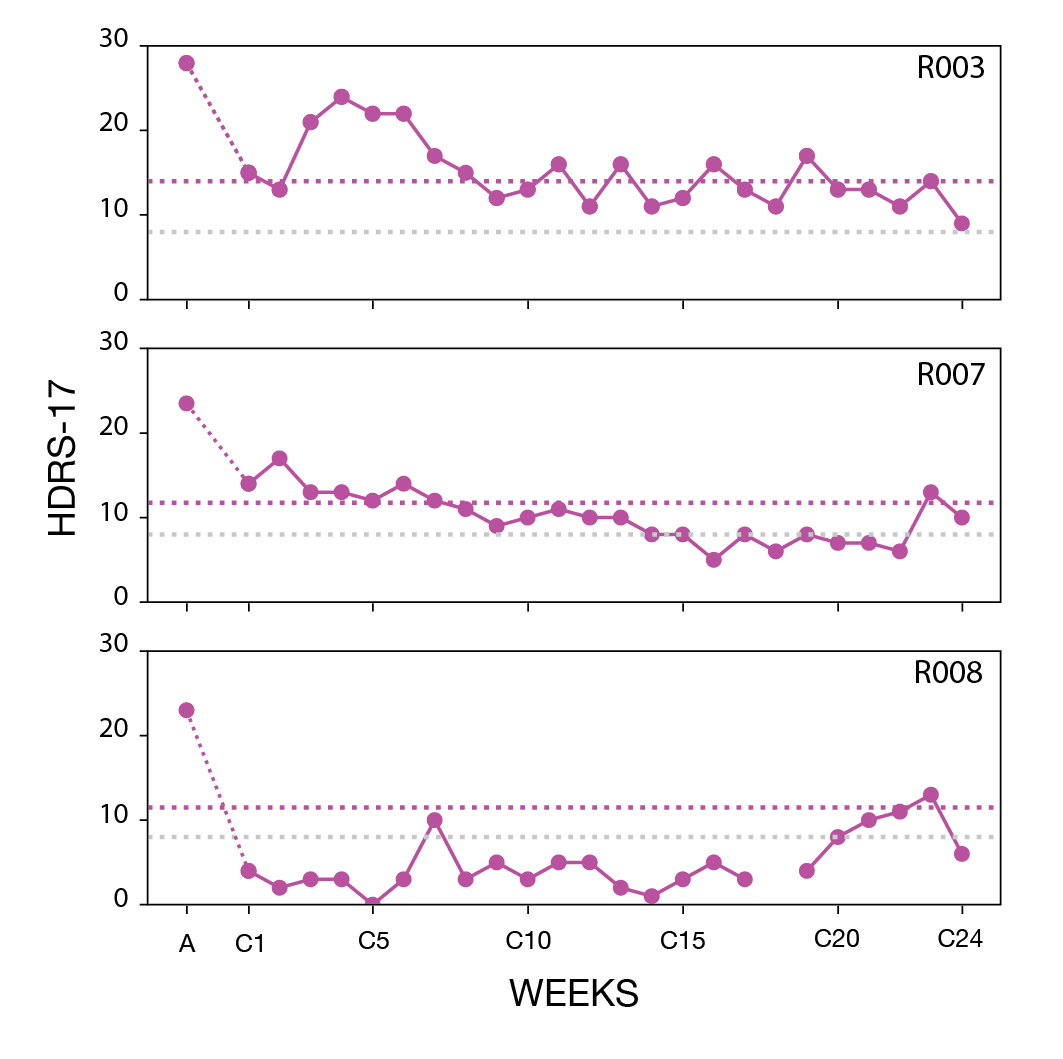

(Purple line): response threshold (Phase A/2) (Grey line): remission rate (8).

**Supp. Fig. 6. SDC and HDRS-17 Trajectories in RC+S**

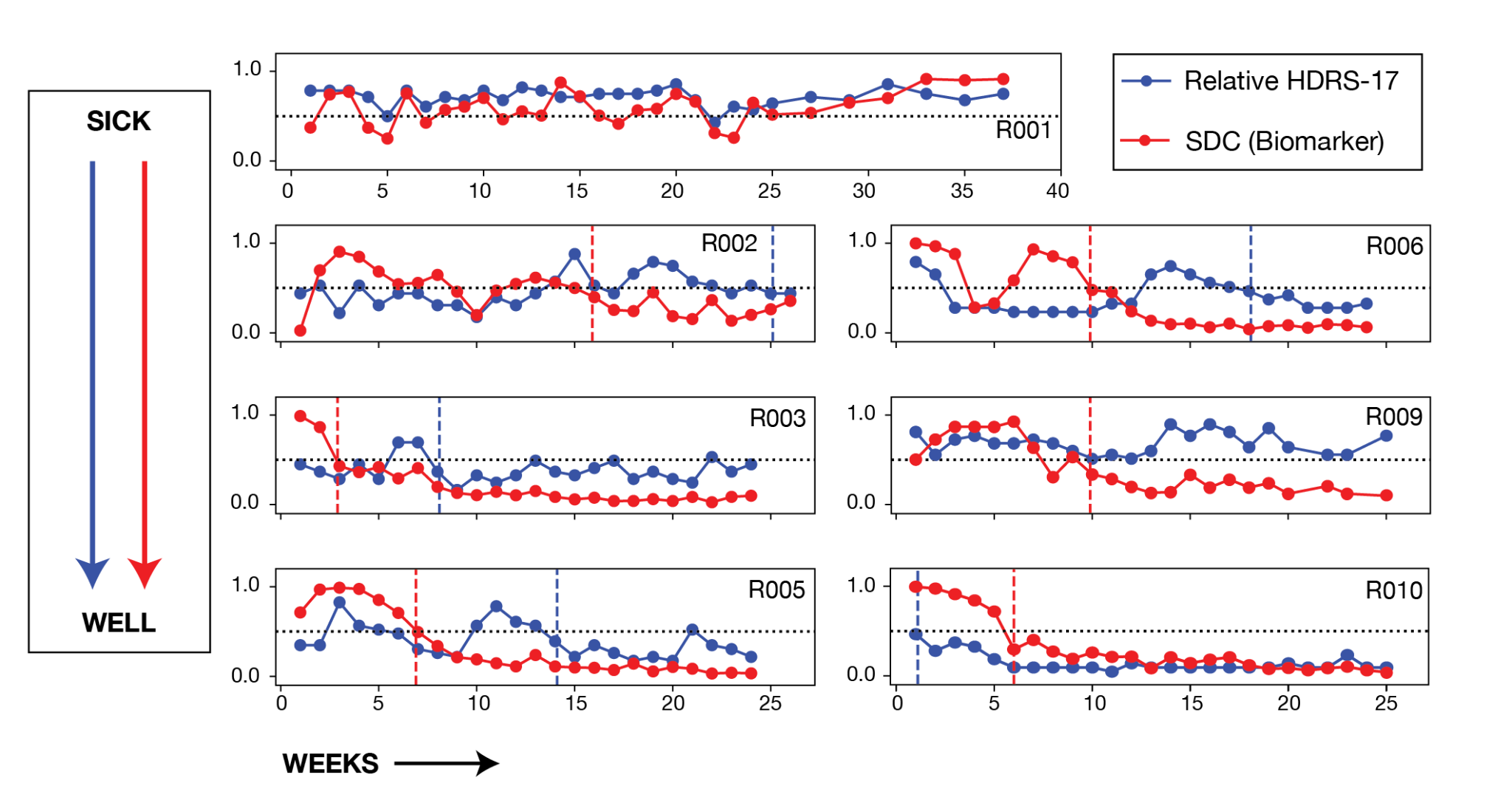

SDC Biomarker and relative HDRS-17 trajectories. Red dashed line refers to the SDC-identified stable response week and the blue dashed line refers to the HDRS-17-identified stable response week. Horizontal black dashed line refers to the threshold of 0.5, used for both measures.

**Supp. Fig. 7. ANX/DEP Bar Plots for SDC/HDRS-17 Incongruent Participants**

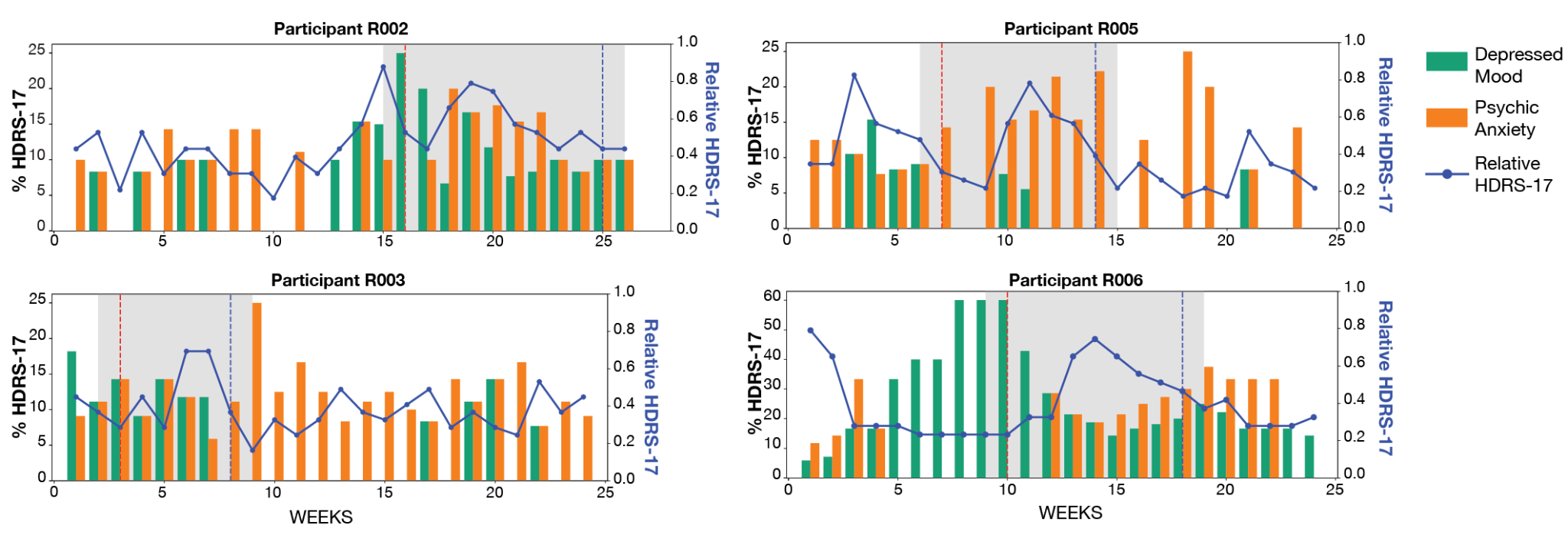

Bar plots showing how Depressed Mood (Teal) and Psychic Anxiety (Orange) items of HDRS-17 contribute to the total HDRS-17 score, with relative HDRS-17 trajectories (compared to baseline) shown for the participants where incongruency is observed with the SDC. The red dashed vertical line shows transition week identified by the SDC and blue dashed vertical line represents HDRS-17-identified transition week. The gray-marked region shows the period in between the transitions. Anxiety symptoms are relatively persistent despite changes in HDRS-17 and mood symptoms (including two subjects showing low mood symptoms by the first week).

**Supp. Fig. 8. STIM ON/OFF Correlations and Stim ON SDC Trajectories**

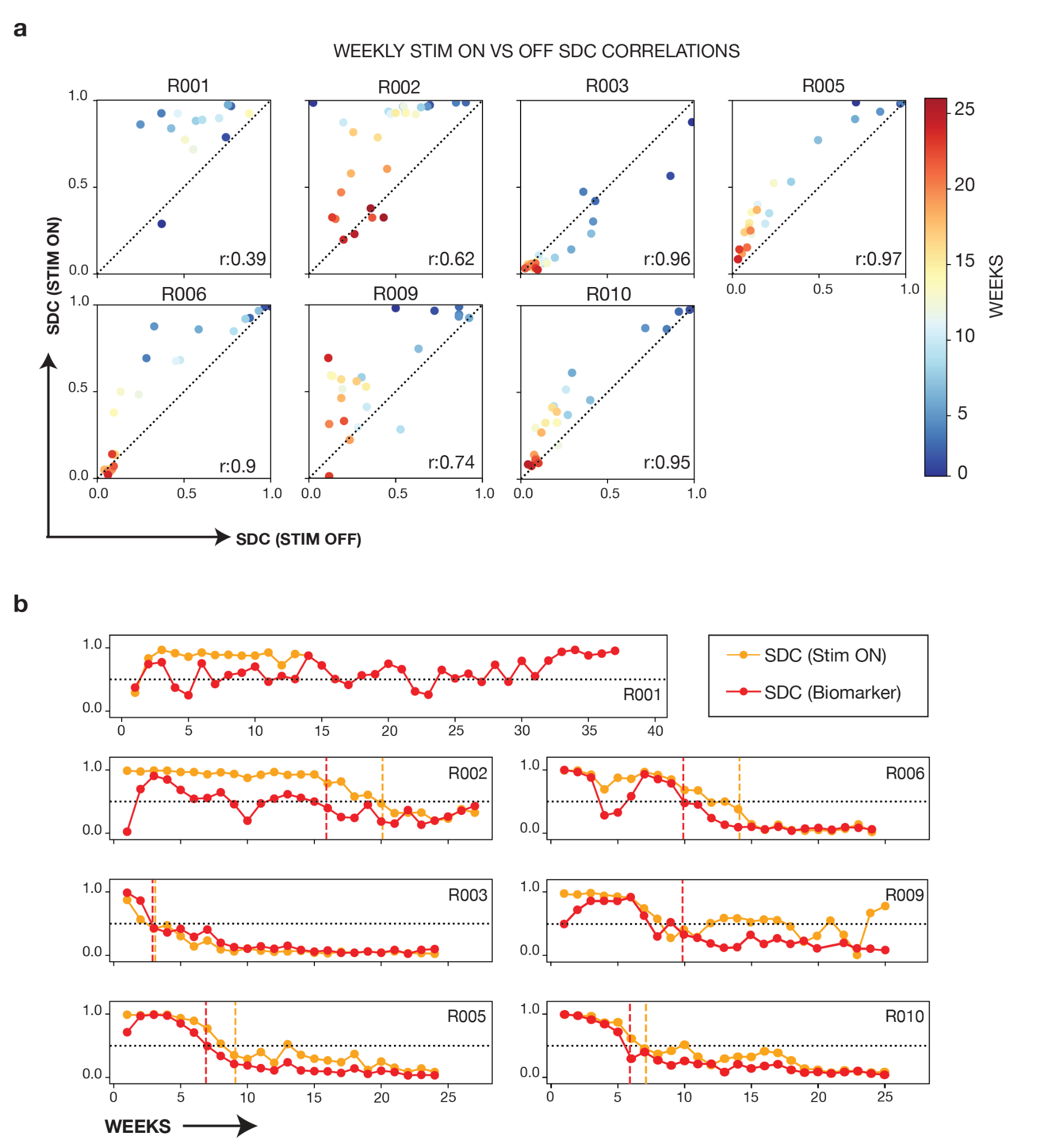

(a) Weekly STIM OFF and STIM ON SDC correlations for each participant.
(b) Illustration of how well the STIM OFF and STIM ON SDC biomarkers match over the observation period for RC+S cohort.

**Supp. Fig. 9. Morning/Evening Correlations**

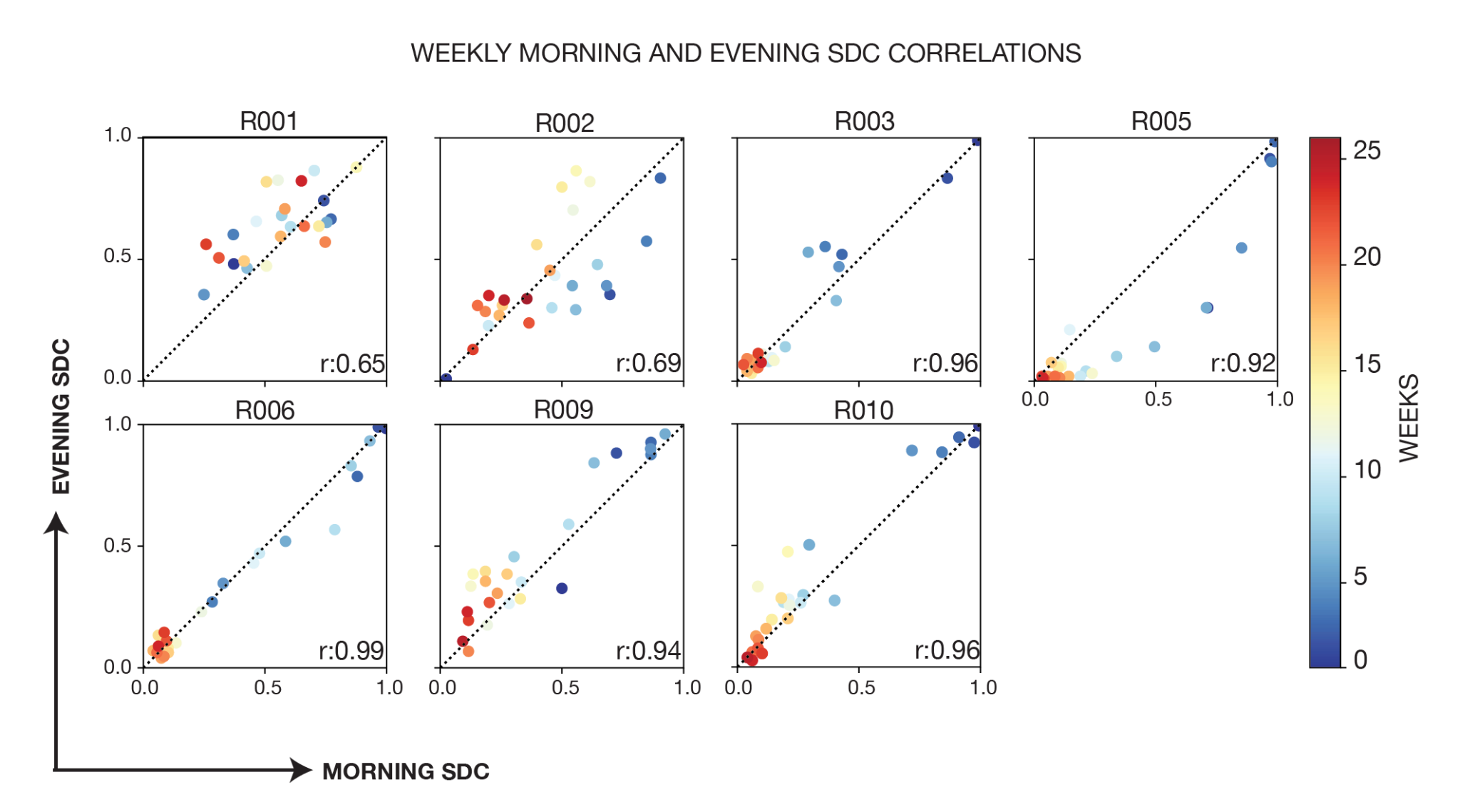

Weekly Morning and Evening SDC correlations for each participant.

**Supp. Fig. 10. Powers and HDRS-17 Trajectories**

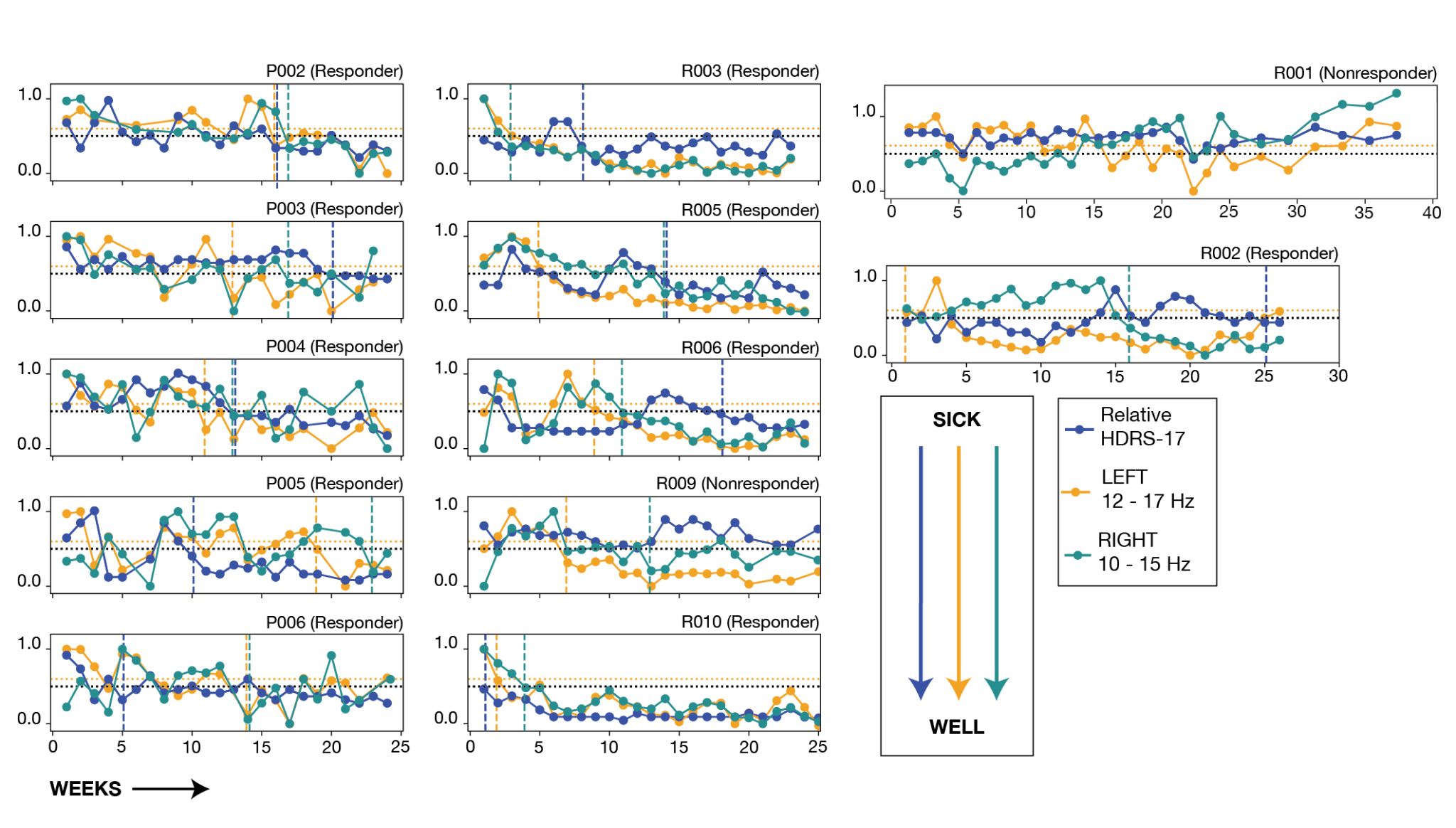

Trajectories of Left-hemisphere (12-17 Hz) and Right-hemisphere (10-15 Hz) powers as surrogates of the SDC, shown with relative HDRS-17 on a common HDRS-17 y-axis (labeled as in Supp. Fig. 6.). The left- and right-hemisphere power trajectories are inverted for display so that, like the relative HDRS-17, lower values correspond to the stable response state. In the underlying data these bands increase with stable recovery, in contrast to the SDC and relative HDRS-17, which decrease; the inversion is for display only and does not change the analysis or the transition thresholds. Vertical blue, gold and teal dashed lines refer to the HDRS-17, left and right identified stable response weeks, respectively. The horizontal dashed lines mark the transition thresholds on the inverted display: 0.5 for the HDRS-17 and right-hemisphere power (blue) and 0.6 for the left-hemisphere power (gold).

**Supp. Fig. 11. Relative HDRS-17 in Responders during Discontinuation**

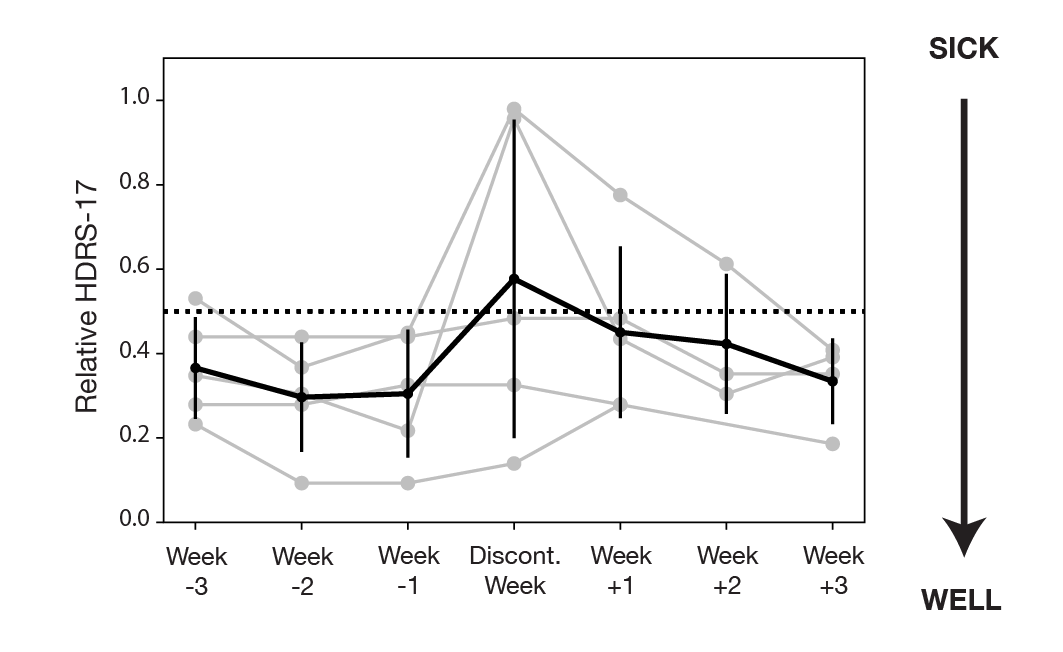

Relative HDRS-17 trajectories around the week of stimulation discontinuation (±3 weeks) in responders. Grey lines show individual responder relative HDRS-17 values; the black line with error bars shows the across-responder mean ± s.d.. Changes around discontinuation are heterogeneous: two participants exhibit a transient increase at the discontinuation week with brief return of symptoms, but relative HDRS-17 subsequently falls back below the 0.5 threshold in the weeks after stimulation is restarted.
